# Modelled mortality benefits of reducing fibrinolysis resistance in sepsis: integrating observational and *ex vivo* data

**DOI:** 10.64898/2026.10.24.26366402

**Authors:** Lucy Coupland, Matthew Self, Eurwin Suryana, Ruby Law, Philip Crispin, Charithani Keragala, Steve Frost, David Rabbolini, Peter K Moore, Robert Medcalf, Anders Aneman

## Abstract

**Background:** Suppressed fibrinolysis (fibrinolysis resistance) is increasingly recognised as a driver of organ failure in sepsis, yet trials in sepsis-induced coagulopathy have largely targeted coagulation rather than fibrinolysis. This study combined an observational ICU cohort with nested analyses of plasma fibrinolysis proteins and *ex vivo* reversibility, modelling the potential mortality reduction from restoring fibrinolytic capacity to inform trial design.

**Methods:** In 188 adults admitted to ICU with sepsis, repeated tissue plasminogen activator (tPA)-enhanced viscoelastometric testing quantified fibrinolysis resistance as the fibrin-adjusted lysis time (TPA-LT/FIBA10) and as ordinal Grades 1–3 of worsening resistance. Adjusted logistic regression and time-varying Cox models related fibrinolysis resistance to death within 28 days of admission, with unmeasured confounding assessed by E-values. In nested sub-studies, plasma fibrinolysis proteins were measured and whole blood was tested ex vivo for reversal of resistance by supplemental tPA, plasminogen, or an inhibitory antibody to α2-antiplasmin (α2AP iAb). Ex vivo effects were modelled onto the observed fibrinolysis resistance–mortality relationship using a hypothetical intervention distribution framework.

**Results:** Each e-fold (∼2.72×) increase in TPA-LT/FIBA10 was associated with a 2.2-fold (95% CI 1.5–3.3) increase in the odds of death in the fully adjusted logistic model and a 2.3-fold (1.6–3.2) increased hazard in the time-varying Cox model. Comparing the least (Grade 1) with the worst (Grade 3) resistance state, the odds ratio was 7.4 (2.4–23) and the hazard ratio 6.7 (3.0–15). Associations were robust to unmeasured confounding, with E-values of 3–13. Grade 3 resistance showed the highest PAI-1/tPA and α2AP/plasminogen ratios. *Ex vivo* supplementation with plasminogen or α2AP iAb reversed fibrinolysis resistance from Grade 3 to Grade 1. Modelling these effects yielded hypothetical absolute mortality reductions of 18-21% across Grades 2 and 3, and 32-36% in Grade 3.

**Conclusions:** Fibrinolysis resistance measured at the bedside in sepsis reflects mechanistically reversible changes in fibrinolytic balance *ex vivo*. Modelling reversal by plasminogen supplementation or α2AP inhibition yielded hypothetical absolute mortality reductions of up to 32% in the most severe resistance state. These hypothesis-generating estimates identify candidate interventions and provide a rationale for a clinical trial.

**Trial registration:** NCT06680180, Registration Date 2024-11-08

## BACKGROUND

The responses of the immune, inflammation and coagulation systems in sepsis are highly interlinked with reciprocal amplification mechanisms that lead to widespread generation of microthrombi [1]. A simultaneous reduction in fibrinolytic activity, referred to as fibrinolysis resistance [2], confers a greater propensity for thrombi to persist and extend [3]. The combination of hypercoagulability and fibrinolysis resistance results in microvascular clotting and obstructive tissue hypoperfusion that may culminate in multiorgan failure and death. Clinical trials in sepsis induced coagulopathy have overwhelmingly investigated anticoagulant strategies and repeatedly returned neutral results in unselected populations [1, 4]. These trials enrolled biologically heterogeneous patients without enrichment for the coagulopathic phenotype most likely to benefit and did not address fibrinolysis resistance [5], which has since been recognised as both dynamic and important to patient-centred outcomes [6–13].

Fibrinolysis resistance in sepsis is associated with reduced levels of the substrate plasminogen and increased levels of plasminogen activator inhibitor type 1 (PAI-1) and α2-antiplasmin (α2AP), the inhibitors of the key fibrinolysis enzymes tissue plasminogen activator (tPA) and plasmin, respectively. However, these mediators are not measured routinely in clinical practice, and assessing them individually fails to reflect the integrated, dynamic interaction among fibrinolytic regulators. Fibrinolytic capacity in whole blood sampled from septic patients can instead be quantified by tPA-enhanced viscoelastometric testing (VET), which can be performed at the bedside and yields results within a clinically actionable timeframe [6, 7]. This point of care assay enables two additional concepts. First, tPA-enhanced VET can be used to monitor the effects of fibrinolytic directed therapy, when administered clinically, and guide dose escalation. Second, whole blood conditions can be manipulated *ex vivo* to estimate therapeutic reversal of fibrinolytic resistance.

A grading system of fibrinolysis resistance in sepsis, using VET-derived lysis time adjusted for the magnitude of fibrin clot, was recently described. Increasing grade of resistance was associated with differences in plasma fibrinolysis proteins, disease severity, and, importantly, requirements for organ support and survival in ICU [6]. This grading system enables sequential and nuanced assessments of fibrinolytic capacity beyond the dichotomous classification of impaired versus normal fibrinolysis. It also captures the dynamic transitions of fibrinolysis resistance needed to identify in which patients and at what time therapeutic benefit might be optimised.

This study used an observational database of patients admitted to ICU with sepsis who underwent multiple tPA-VET assays from admission onwards, in whom the prognostic association between fibrinolysis resistance and mortality was quantified, including the robustness to unmeasured confounding. In nested sub-studies, plasma fibrinolysis proteins were measured to characterise the fibrinolytic balance underlying the resistance grades, and fresh whole blood was tested for the capacity of supplemental tPA, plasminogen, or an inhibitory α2AP antibody (α2AP iAb) added *ex vivo* to restore fibrinolytic capacity. We hypothesised that modelling these *ex vivo* effects onto the observed exposure-response relationship between fibrinolysis resistance and death within a hypothetical interventional distribution framework, could estimate the magnitude of potential treatment effects. This approach aligns with the PROGRESS 4 stratified-medicine framework [14] and could inform the design of a future interventional trial.

## METHODS

This study was performed in Liverpool Hospital ICU, Sydney, Australia within a research program approved by the South Western Sydney Local Health District Human Research Ethics Committee (2022/ETH02122). Consent was waived as permitted under the National Statement on Ethical Conduct in Human Research 2023 for low-risk research in critically ill patients. Patients were recruited and VET data collected prospectively, with de-identified clinical and pathology data added retrospectively, as part of the FORTITUDE research program on fibrinolysis resistance (NCT06680180). The study adheres to the STROBE guidelines for observational studies [15] (see Table S1 in Additional File 1).

### Participants

Adult patients (≥18 years) admitted to ICU between January 2023 and April 2026 were screened and included if diagnosed with sepsis according to Sepsis-3 criteria [16] from a suspected or confirmed infection, were for full active management and expected to survive beyond the day following admission, and staff were available to perform the first VET within 24 hours of admission. Patients with a pre-existing chronic coagulation disorder, known pregnancy, admitted with palliative intent or with the first VET performed after 24 hours of admission were excluded.

### Clinical data and outcome variables

Study data included patient characteristics, APACHE III (Acute Physiology and Chronic Health Evaluation), SOFA (Sequential Organ Failure Assessment), SIC (Sepsis Induced Coagulopathy) [17], and DIC (disseminated intravascular coagulation) [18] scores, clinical frailty score [19], standard blood test results, organ support measures, therapeutic anticoagulant and steroid administration, and survival up to 28 days after ICU admission. The study design considered feedback from patient surviving sepsis (see Acknowledgement).

### Viscoelastometric analyses and fibrinolysis assessment

The ClotPro® (Haemoview Diagnostics, Milton, QLD, Australia) VET point-of-care platform was used to assess coagulation and fibrinolysis. Blood was collected into 3.2% buffered sodium citrate with three assays performed within 30 minutes: (i) EX-test - whole blood coagulation activated with tissue factor with a heparin inhibitor included; (ii) FIB-test - same as the EX-test with two platelet inhibitors included to assess fibrin clot formation; and (iii) TPA-test - same as the EX-test with tPA (650 ng/mL) added to permit evaluation of fibrinolysis. The VET parameters reported include: EX-test clotting time, the time for the forming clot to reach 2 mm amplitude (EX-CT, sec); the FIB- and EX-test clot amplitudes 10 min after commencement of clot formation (FIB or EXA10, mm); the TPA-test clot lysis time, the time taken for 50% of the maximum clot firmness to be lysed (TPA-LT, sec), TPA-test maximum clot firmness/amplitude measured during the 60-minute assay run time (TPA MCF, mm). A fibrin-adjusted measure of fibrinolysis was obtained by dividing the TPA-LT by the FIBA10 and referred to as TPA-LT/FIBA10 (sec/mm) [6]. Distinct grades (Grades 1-3) of fibrinolysis resistance were defined by the TPA-LT/FIBA10 variable, with a higher grade reflecting increasing resistance, as previously described [6]. The VET assay schedule included days 1, 2, 3, 5, 7, 10 and 15 after admission to ICU, for as long as the patient remained in ICU.

### Detection of plasma fibrinolysis proteins

In a nested sub-study, blood collected in 3.2% buffered sodium citrate at the time of the VET assays was centrifuged at 5000 x g for 20 min at room temperature to obtain platelet-poor plasma and frozen at − 80°C. ELISA kits (Innovative Research, Novi, MI, USA) were used to measure tPA activity (IHUTPAKT), PAI-1 activity (IHUPAI1KT), plasminogen (IHUPLGKT) and α2AP (IHUA2APLKTT). Plasma samples were diluted and processed following the manufacturer’s instructions. The log-scale ratios of PAI-1/tPA and α2AP/plasminogen are reported to summarise the fibrinolysis balance.

### Reversibility of fibrinolysis resistance ex vivo

In a second nested sub-study, the reversibility of fibrinolysis resistance was tested *ex vivo* in whole blood concurrent to the VET assessments focusing on patients with Grade 2 and Grade 3 fibrinolysis resistance. The standard TPA-(650 ng/mL (∼10nM) tPA) test was supplemented with the following agents by addition to the analytical cup immediately prior to performing the assay: i) alteplase (tPA; 650 ng/mL; Boehringer-Ingelheim, Ingelheim, Germany) thus doubling the tPA dose to a total of 1.3 μg/mL in the TPA-test; ii) plasminogen (143 μg/mL; Hyphen Biomed, Neuville-sur-Oise, France); iii) an inhibitory α2AP antibody (α2AP iAb; 124 μg/mL equivalent to 5 μmol/L, Prof Ruby Law, Monash University, Melbourne, Australia).

### Statistical methods

No formal sample size or power calculations were performed given the exploratory nature of this study. Patient characteristics, ICU interventions and coagulation variables are reported as medians [interquartile range, IQR] as appropriate for non-normally distributed data. The primary outcome was death, with discharge alive treated as a competing event, within 28 days of admission to ICU. The cross-sectional association between admission fibrinolysis and ICU mortality was examined by binary logistic regression using a primary continuous exposure (TPA-LT/FIBA10, natural log-transformed) as well as an ordinal exposure (fibrinolysis resistance Grade 1-3). Data-driven definitions of fibrinolysis resistance grades were obtained by unsupervised, univariate cluster analysis using a Gaussian Mixture Model with Box-Cox transformed data as previously reported for 116 patients [6]. In addition to an unadjusted logistic regression model, minimally adjusted (age, sex), and fully adjusted (age, sex, use of therapeutic anticoagulation, SOFA score contemporaneous to each VET assay, the time for each assay defined as hours since admission to ICU) models were specified. The latter was further adjusted to include the use of steroids (hydrocortisone ≥200 mg, or dexamethasone >8 mg, or methylprednisolone >500 mg) since glucocorticoids are reported to increase PAI-1 levels in inflammatory states [20, 21]. No imputations were made for missing data in the adjusted models that are reported with the number of patients included in each specified.

The exposure-response relationship between TPA-LT/FIBA10 and mortality was studied using restricted cubic splines with three knots placed at the 10th, 50th, and 90th percentiles of log(TPA-LT/FIBA10) [22], with non-linearity assessed by the likelihood ratio test. Competing risks were modelled using the Fine-Gray sub-distribution hazard and cause-specific Cox regression. The proportional hazards assumption was assessed using Schoenfeld residuals. Violation was anticipated because both SOFA and the prognostic importance of fibrinolysis resistance change dynamically over time [6] and was addressed by adding a log(time) interaction term. Time-varying associations were modelled in counting process format with start-stop intervals defined by the times between VET assays. Robust sandwich standard errors were used to account for within-patient correlation across serial measurements. Unmeasured confounding in the observed associations between fibrinolysis resistance and patient outcomes was examined by computing E-values for the risk ratio effect estimate including its 95% confidence interval (95%CI) [23, 24]. The E-value represents the minimum risk ratio that an unmeasured confounder would need to have for both the exposure and the outcome to fully explain away the observed association after adjustment for measured covariates.

Differences in the balance of key fibrinolysis proteins across resistance grades were analysed using a linear mixed-effects model with fibrinolysis resistance grade as a fixed effect and a random intercept for patient to account for within-subject correlation. *Ex vivo* reversibility by adding tPA, plasminogen or an α2AP iAb was summarised as median [IQR] post/pre-supplementation TPA-LT/FIBA10 ratios within each resistance grade and compared across grades using Mann-Whitney U and Kruskal-Wallis tests.

To translate the *ex vivo* fibrinolysis effects into mortality estimates on the scale of the observational cohort, a hypothetical interventional distribution framework was used [25–27], see schematic Figure 1. Each patient’s observed TPA-LT/FIBA10 was shifted by the ratio of the post- to pre-TPA-LT/FIBA10 measured *ex vivo* following supplemental tPA, plasminogen or the α2AP iAb. A ratio = 1 indicates no effect with decreasing values <1 indicating increased fibrinolysis. The ratio was applied to the observational exposure-response model via logit-scale regression using bootstrapping with 1000 iterations. The counterfactual mortality was derived from the fully adjusted logistic exposure-response model. The code used for analyses is available at https://zenodo.org/records/22821116.

**Figure 1.**
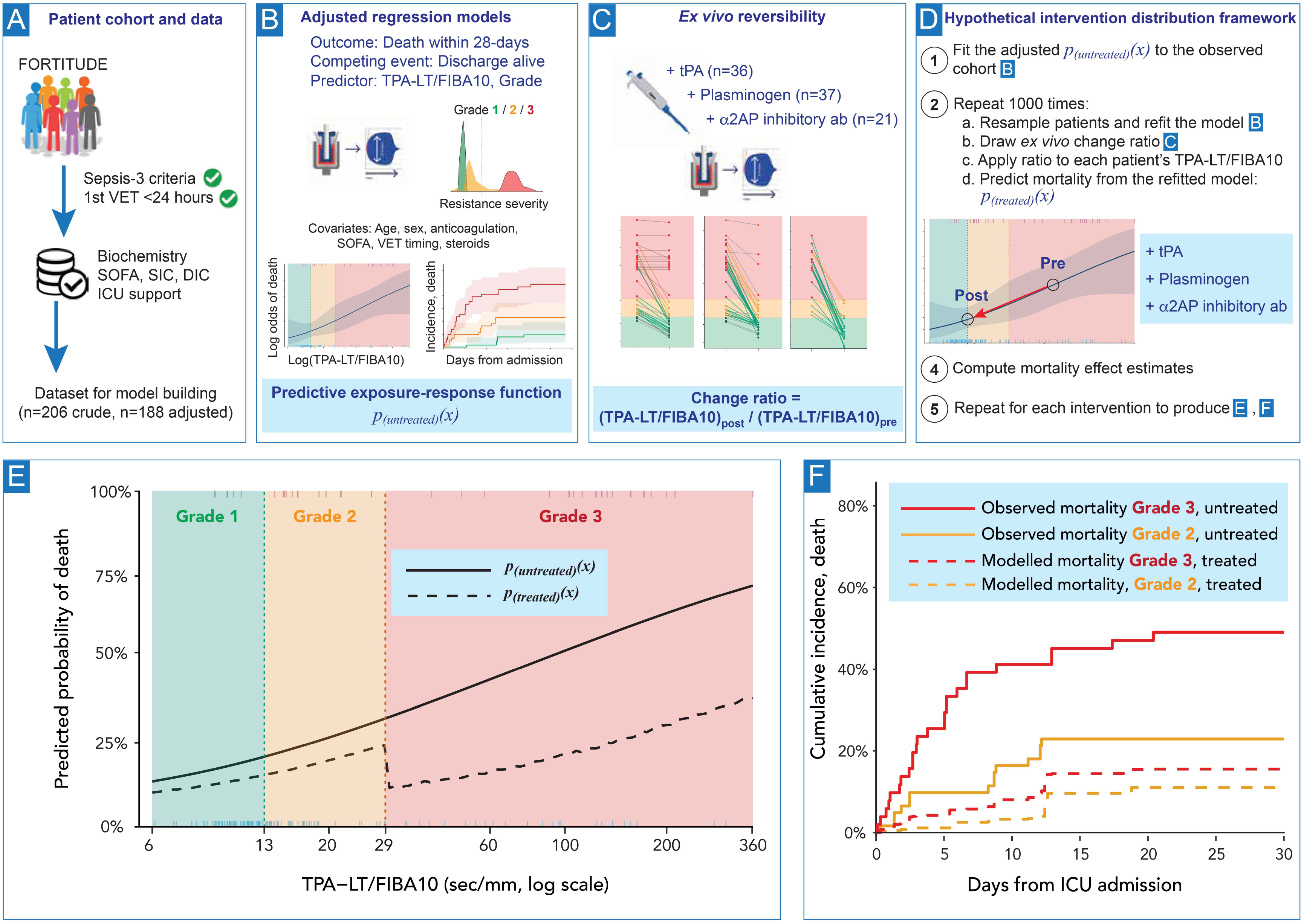
Schematic of the hypothetical interventional distribution framework [25–27]. **(A)** Patients with sepsis meeting Sepsis-3 criteria and undergoing a first viscoelastometric test (VET) within 24 hours of ICU admission formed the analytic cohort (full cohort n=206; adjusted models n=188). **(B)** Adjusted regression models related fibrinolysis resistance, as the fibrin adjusted lysis time (TPA-LT/FIBA10) and as ordinal Grades 1-3, to death within 28 days, with discharge alive as a competing event. An exposure-response function *p(_untreated_)(x)* and a competing-risks model with cumulative incidence function were produced. **(C)** In separate *ex vivo* experiments, paired whole-blood samples were tested before and after addition of tPA, plasminogen or an inhibitory α2AP antibody, giving a within-sample change ratio (post/pre TPA-LT/FIBA10). **(D)** The framework combines (B) and (C). Each patient’s observed fibrinolysis resistance is multiplied by a change ratio drawn from *ex vivo* experiments of the same grade, shifting that patient leftward along the exposure-response curve (Pre → Post), and their counterfactual mortality is read from the model. Uncertainty is obtained by a nested bootstrap of 1000 iterations in which patients are resampled and the model refitted, and change ratios are resampled within grade, so that observational variability and uncertainty in the *ex vivo* effect are propagated jointly. **(E)** Modelled predicted probability of death against log(TPA-LT/FIBA10), with *p(_untreated_)(x)* represented by the solid line and *p(_treated_)(x)* represented by the dashed line. **(F)** Observed (solid line) and modelled (dashed line) cumulative incidence of death for in Grade 2 (orange) and Grade 3 (red) fibrinolysis resistance states.

Statistical significance was set at a two-sided p-value of <0.05. All statistical analyses were performed using R 4.3.3 (R Core Team, 2024) with R scripts written using assistance from Claude (Sonnet 4.6, Anthropic 2025, https://www.anthropic.com) and with all outputs reviewed by the authors for interpretation. An interactive web application built with R Shiny using the observational dataset associating fibrinolysis resistance and mortality is available at https://fortitude.shinyapps.io/FORTITUDESHINY/ to demonstrate the impact of a wide range of hypothetical intervention effects to restore fibrinolysis capacity.

## RESULTS

### Participants and clinical characteristics

A total of 325 patients with sepsis or septic shock were admitted to ICU during the study period with 206 patients meeting all inclusion and no exclusion criteria and staff available to perform the first VET, thus representing an observational cohort almost double that previously reported [6]. Septic shock was present in 114 (55%) patients. Patient characteristics, illness severity, organ support measures, source of infection, routine haematology and VET tests are reported in Table 1. Patients who died or demonstrated the most severe fibrinolysis resistance (Grade 3) had greater disease severity scores with increased need for organ support and showed a consumptive coagulopathy. Seven (3%) patients were co-infected with SARS-CoV-2. Respiratory infections dominated (29%) followed by abdominal (19%), soft tissue (13%), and urological (11%) while 28% had combined or other sources of infection. Details of microbiological isolates are provided in the Additional File 1, Table S2. The median time from ICU admission to the first VET was 6.9 hours (IQR 3.2-13) with 3 (IQR 2-6) sequential VET performed per patient separated by 25 (IQR 23-48) hours (VET assay time details shown in Additional File 1, Figure S1).

**Table 1.**
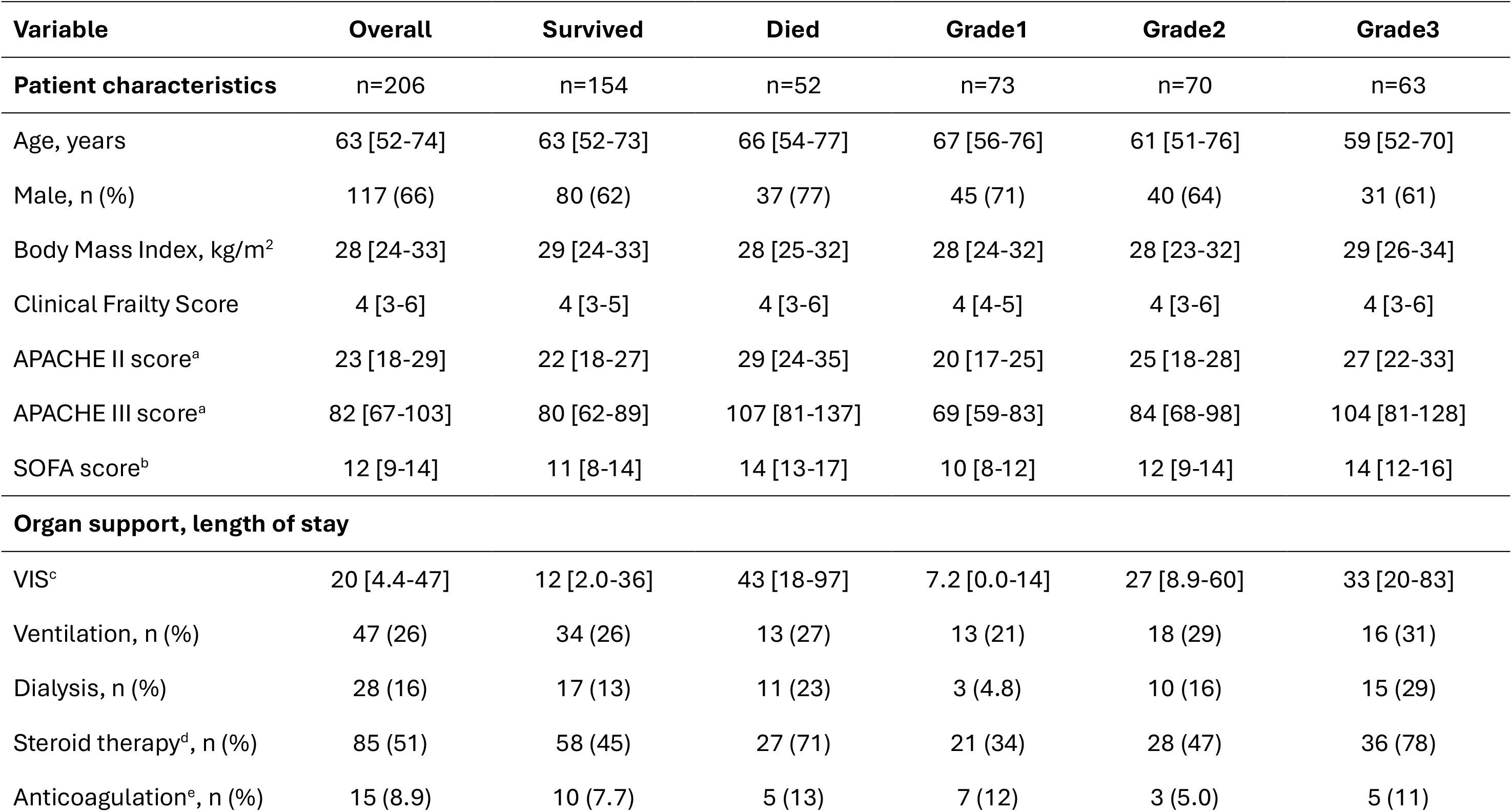
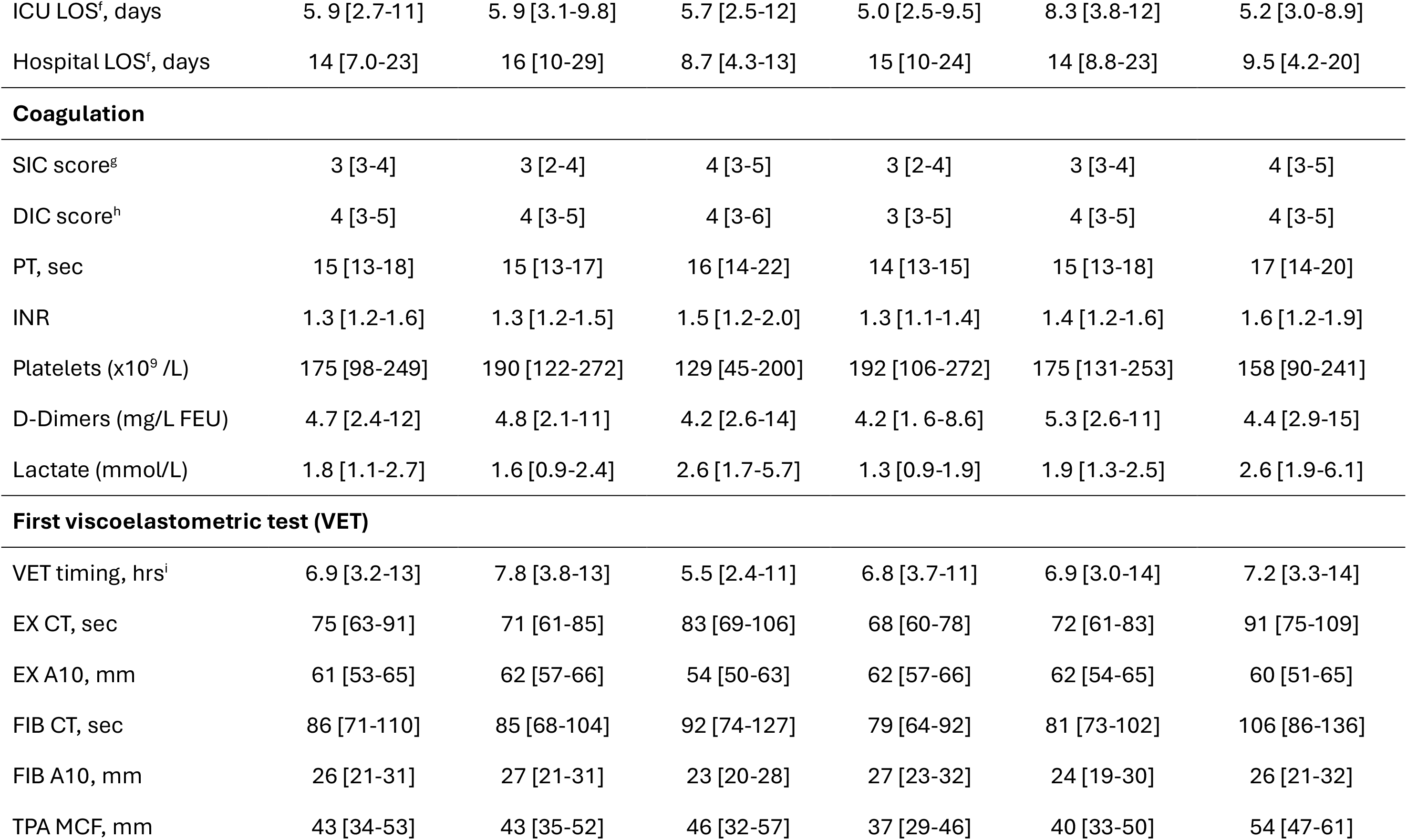
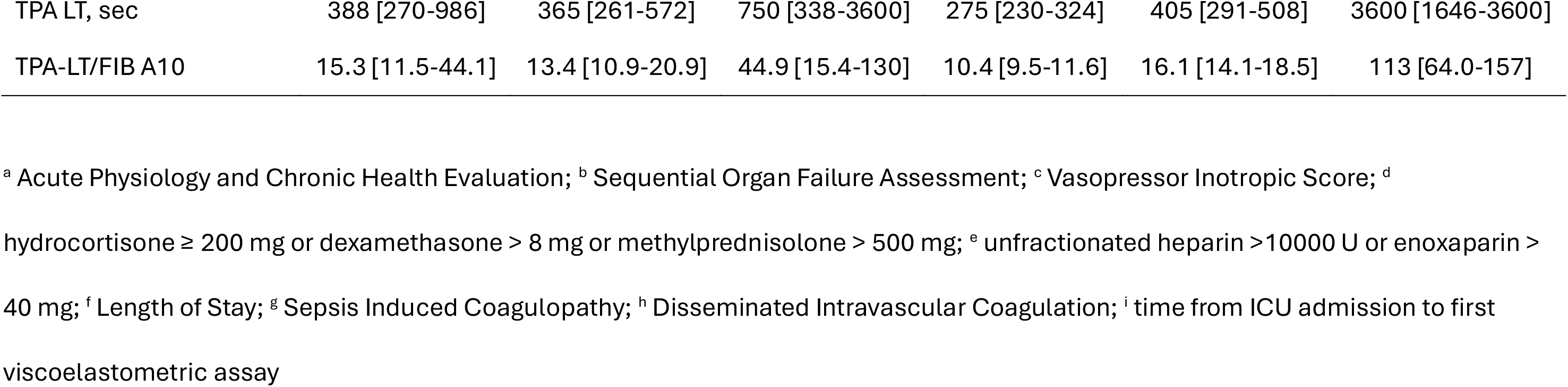
Study cohort characteristics on admission to ICU including standard coagulation and viscoelastometric variables.

| Variable | Overall | Survived | Died | Grade1 | Grade2 | Grade3 |
| --- | --- | --- | --- | --- | --- | --- |
| Patient characteristics | n=206 | n=154 | n=52 | n=73 | n=70 | n=63 |
| Age, years | 63 [52-74] | 63 [52-73] | 66 [54-77] | 67 [56-76] | 61 [51-76] | 59 [52-70] |
| Male, n (%) | 117 (66) | 80 (62) | 37 (77) | 45 (71) | 40 (64) | 31 (61) |
| Body Mass Index, kg/m<sup>2</sup> | 28 [24-33] | 29 [24-33] | 28 [25-32] | 28 [24-32] | 28 [23-32] | 29 [26-34] |
| Clinical Frailty Score | 4 [3-6] | 4 [3-5] | 4 [3-6] | 4 [4-5] | 4 [3-6] | 4 [3-6] |
| APACHE II score<sup>a</sup> | 23 [18-29] | 22 [18-27] | 29 [24-35] | 20 [17-25] | 25 [18-28] | 27 [22-33] |
| APACHE III score<sup>a</sup> | 82 [67-103] | 80 [62-89] | 107 [81-137] | 69 [59-83] | 84 [68-98] | 104 [81-128] |
| SOFA score<sup>b</sup> | 12 [9-14] | 11 [8-14] | 14 [13-17] | 10 [8-12] | 12 [9-14] | 14 [12-16] |
| Organ support, length of stay |  |  |  |  |  |  |
| VIS<sup>c</sup> | 20 [4.4-47] | 12 [2.0-36] | 43 [18-97] | 7.2 [0.0-14] | 27 [8.9-60] | 33 [20-83] |
| Ventilation, n (%) | 47 (26) | 34 (26) | 13 (27) | 13 (21) | 18 (29) | 16 (31) |
| Dialysis, n (%) | 28 (16) | 17 (13) | 11 (23) | 3 (4.8) | 10 (16) | 15 (29) |
| Steroid therapy<sup>d</sup>, n (%) | 85 (51) | 58 (45) | 27 (71) | 21 (34) | 28 (47) | 36 (78) |
| Anticoagulation<sup>e</sup>, n (%) | 15 (8.9) | 10 (7.7) | 5 (13) | 7 (12) | 3 (5.0) | 5 (11) |
| ICU LOS<sup>f</sup>, days | 5.9 [2.7-11] | 5.9 [3.1-9.8] | 5.7 [2.5-12] | 5.0 [2.5-9.5] | 8.3 [3.8-12] | 5.2 [3.0-8.9] |
| Hospital LOS<sup>f</sup>, days | 14 [7.0-23] | 16 [10-29] | 8.7 [4.3-13] | 15 [10-24] | 14 [8.8-23] | 9.5 [4.2-20] |
| Coagulation |  |  |  |  |  |  |
| SIC score<sup>g</sup> | 3 [3-4] | 3 [2-4] | 4 [3-5] | 3 [2-4] | 3 [3-4] | 4 [3-5] |
| DIC score<sup>h</sup> | 4 [3-5] | 4 [3-5] | 4 [3-6] | 3 [3-5] | 4 [3-5] | 4 [3-5] |
| PT, sec | 15 [13-18] | 15 [13-17] | 16 [14-22] | 14 [13-15] | 15 [13-18] | 17 [14-20] |
| INR | 1.3 [1.2-1.6] | 1.3 [1.2-1.5] | 1.5 [1.2-2.0] | 1.3 [1.1-1.4] | 1.4 [1.2-1.6] | 1.6 [1.2-1.9] |
| Platelets (x10<sup>9</sup> /L) | 175 [98-249] | 190 [122-272] | 129 [45-200] | 192 [106-272] | 175 [131-253] | 158 [90-241] |
| D-Dimers (mg/L FEU) | 4.7 [2.4-12] | 4.8 [2.1-11] | 4.2 [2.6-14] | 4.2 [1.6-8.6] | 5.3 [2.6-11] | 4.4 [2.9-15] |
| Lactate (mmol/L) | 1.8 [1.1-2.7] | 1.6 [0.9-2.4] | 2.6 [1.7-5.7] | 1.3 [0.9-1.9] | 1.9 [1.3-2.5] | 2.6 [1.9-6.1] |
| First viscoelastometric test (VET) |  |  |  |  |  |  |
| VET timing, hrs<sup>i</sup> | 6.9 [3.2-13] | 7.8 [3.8-13] | 5.5 [2.4-11] | 6.8 [3.7-11] | 6.9 [3.0-14] | 7.2 [3.3-14] |
| EX CT, sec | 75 [63-91] | 71 [61-85] | 83 [69-106] | 68 [60-78] | 72 [61-83] | 91 [75-109] |
| EX A10, mm | 61 [53-65] | 62 [57-66] | 54 [50-63] | 62 [57-66] | 62 [54-65] | 60 [51-65] |
| FIB CT, sec | 86 [71-110] | 85 [68-104] | 92 [74-127] | 79 [64-92] | 81 [73-102] | 106 [86-136] |
| FIB A10, mm | 26 [21-31] | 27 [21-31] | 23 [20-28] | 27 [23-32] | 24 [19-30] | 26 [21-32] |
| TPA MCF, mm | 43 [34-53] | 43 [35-52] | 46 [32-57] | 37 [29-46] | 40 [33-50] | 54 [47-61] |
| TPA LT, sec | 388 [270-986] | 365 [261-572] | 750 [338-3600] | 275 [230-324] | 405 [291-508] | 3600 [1646-3600] |
| TPA-LT/FIB A10 | 15.3 [11.5-44.1] | 13.4 [10.9-20.9] | 44.9 [15.4-130] | 10.4 [9.5-11.6] | 16.1 [14.1-18.5] | 113 [64.0-157] |

<sup>a</sup> Acute Physiology and Chronic Health Evaluation; <sup>b</sup> Sequential Organ Failure Assessment; <sup>c</sup> Vasopressor Inotropic Score; <sup>d</sup>
hydrocortisone ≥ 200 mg or dexamethasone $>$ 8 mg or methylprednisolone $>$ 500 mg; <sup>e</sup> unfractionated heparin $>$ 10000 U or enoxaparin $>$
40 mg; <sup>f</sup> Length of Stay; <sup>g</sup> Sepsis Induced Coagulopathy; <sup>h</sup> Disseminated Intravascular Coagulation; <sup>i</sup> time from ICU admission to first
viscoelastometric assay

### Fibrinolysis assessments and outcome

The cross-sectional logistic regression model for the associations between admission log(TPA-LT/FIBA10) or fibrinolysis resistance grades and death is reported in Table 2 together with the time-varying Cox analysis including the dynamic, repeated measures. Co-variates were missing for 18 patients with 188 patients (711 VET assays) included in the adjusted cohort. Admission disease acuity, fibrinolysis resistance and outcomes were not significantly different in the excluded patients (see Additional File 1, Table S3) compared to the retained cohort. The crude, minimally adjusted, and fully adjusted regression models showed broadly similar associations, with no meaningful change in direction and only modest changes in magnitude. This supports that fibrinolysis resistance was associated with increased risk of death independent of age, sex, anticoagulation, contemporaneous SOFA score, time since ICU admission and use of steroids. Each e-fold (∼2.72) increase in log(TPA-LT/FIBA10) was associated with at least a doubling of the odds of death.

**Table 2.**
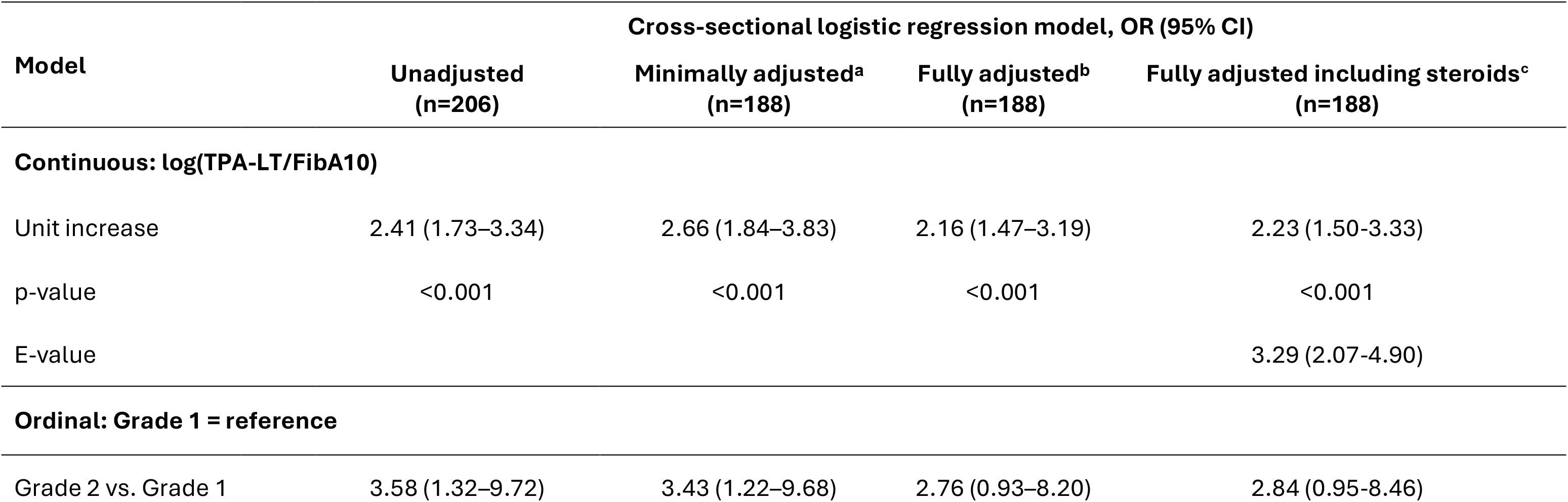
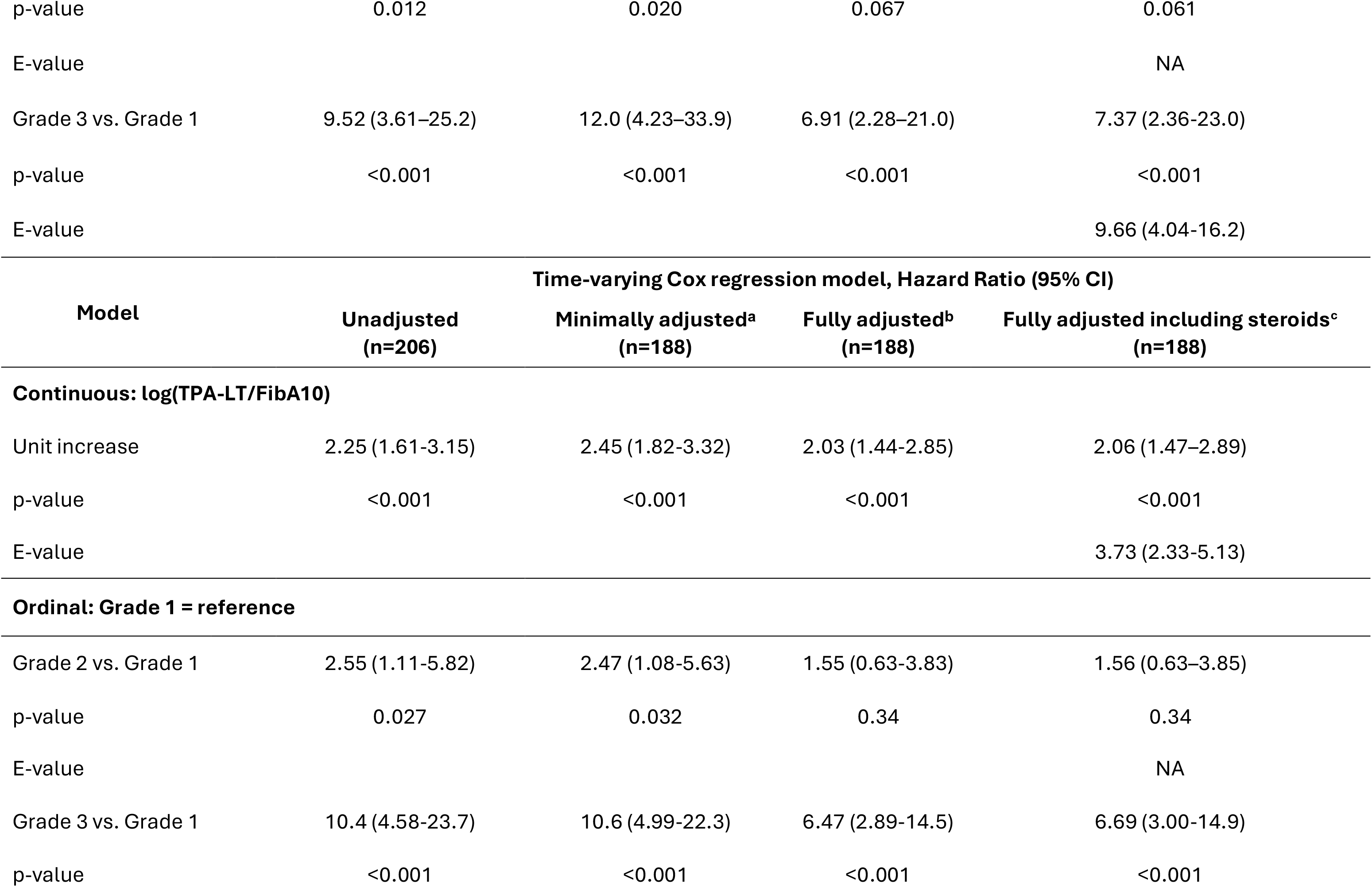
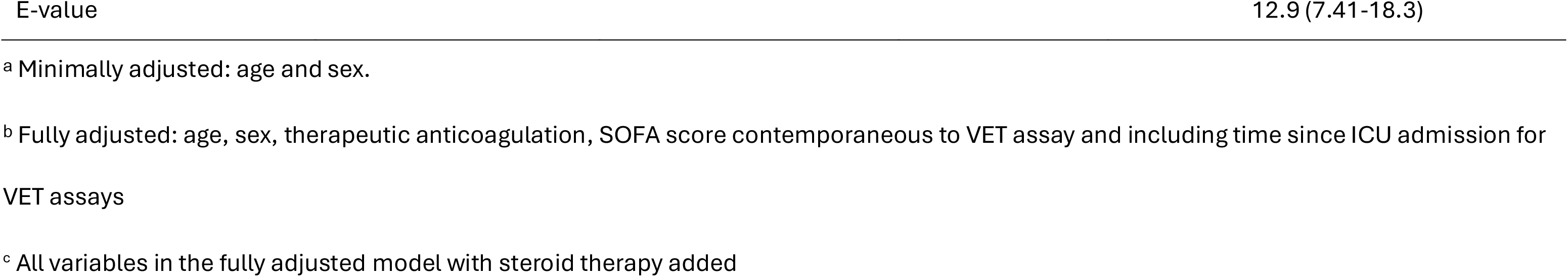
Associations between fibrinolysis resistance and mortality. The cross-sectional logistic regression model used both the continuous (logTPA-LT/FibA10) as well as the ordinal measurements (Grade) of fibrinolysis resistance on admission to ICU. The odds ratios (OR) are reported including the 95% confidence intervals (95%CI) for the unadjusted and the adjusted models. The OR for log(TPA-LT/FibA10) is reported per 1-unit increase equivalent to e-fold (∼2.72×) increase in the raw TPA-LT/FIBA10 value. Since the SOFA score and TPA-LT/FIBA10 fibrinolysis resistance variable changed dynamically across repeated measures, the hazard ratios (HR) from a time-varying Cox regression model are included. The robustness of the underlying fibrinolysis resistance–mortality associations to unmeasured confounding in both fully adjusted models are reported by the E-value. The E-value is the minimum strength of association, on the risk-ratio scale, that an unmeasured confounder would need with both the exposure and mortality to fully explain away the estimate.

The clustering analysis defined Grade 1 as a TPA-LT/FIBA10 <13 sec/mm, Grade 2 as ≥13 and <29 sec/mm and Grade 3 as ≥29 sec/mm, thus were very similar to those previously reported in a smaller sepsis cohort [6] (see Additional File 1, Figure S2). Relative to Grade 1, Grade 2 fibrinolysis resistance lost significance in the adjusted models, whereas Grade 3 was associated with a 7- to 12-fold increased risk of death in the cross-sectional model. The results were similar using the time-varying hazard ratios. The significant observed associations between fibrinolysis resistance and mortality in the fully adjusted models were strongly (E-value >3) and very strongly (E-value >9) robust to unmeasured confounding (Table 2) for the continuous log(TPA-LT/FIBA10) and the Grade 3 vs Grade 1 comparisons, respectively.

The relationship between log(TPA-LT/FIBA10) and the predicted probability of death is shown in Figure 2. The probability of death increased from 19 (11-31)% at the Grade 1/2 transition to 31 (15-52)% at the Grade 2/3 transition. The time-varying relationships between fibrinolysis grades and the cumulative incidence of death or discharge alive are shown in Figure 3, demonstrating that Grade 3 resistance was associated with a higher incidence of death (top) and lower incidence of being discharged alive (bottom) from ICU across all time points.

**Figure 2.**
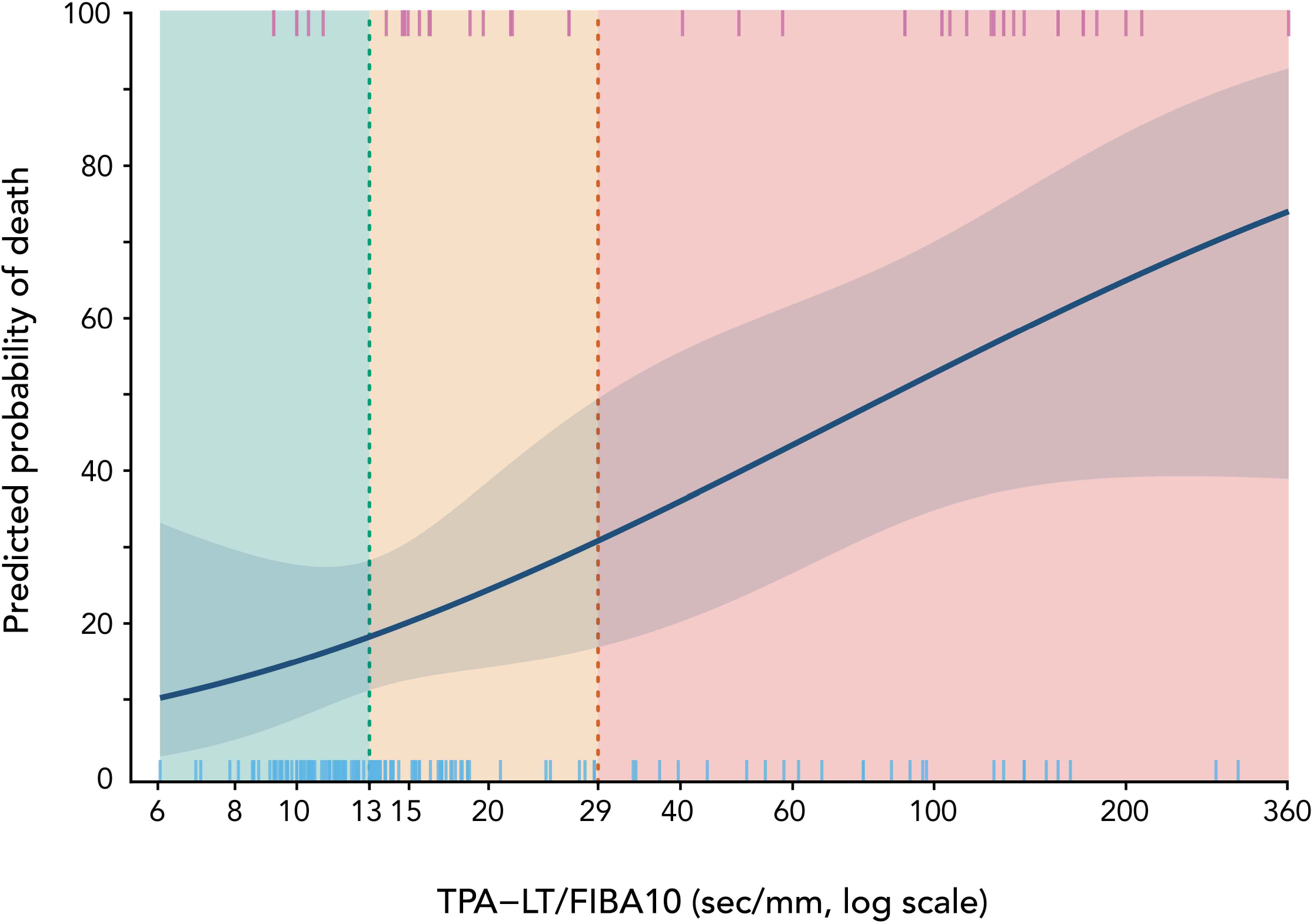
The relationship between admission fibrinolysis resistance (continuous TPA-LT/FIBA10 on the log scale) and predicted mortality up to 28 days. The solid line is the cubic spline logistic regression of the fully adjusted model (age, sex, anticoagulation, SOFA, time since admission to ICU for VET and steroids) with the shaded area representing its 95% confidence interval. The rug marks represent survival (blue marks bottom) and death (red marks top). The dashed vertical lines represent the thresholds between the ordinal Grade 1 (green), Grade 2 (yellow) and Grade 3 (red) fibrinolysis resistance states.

**Figure 3.**
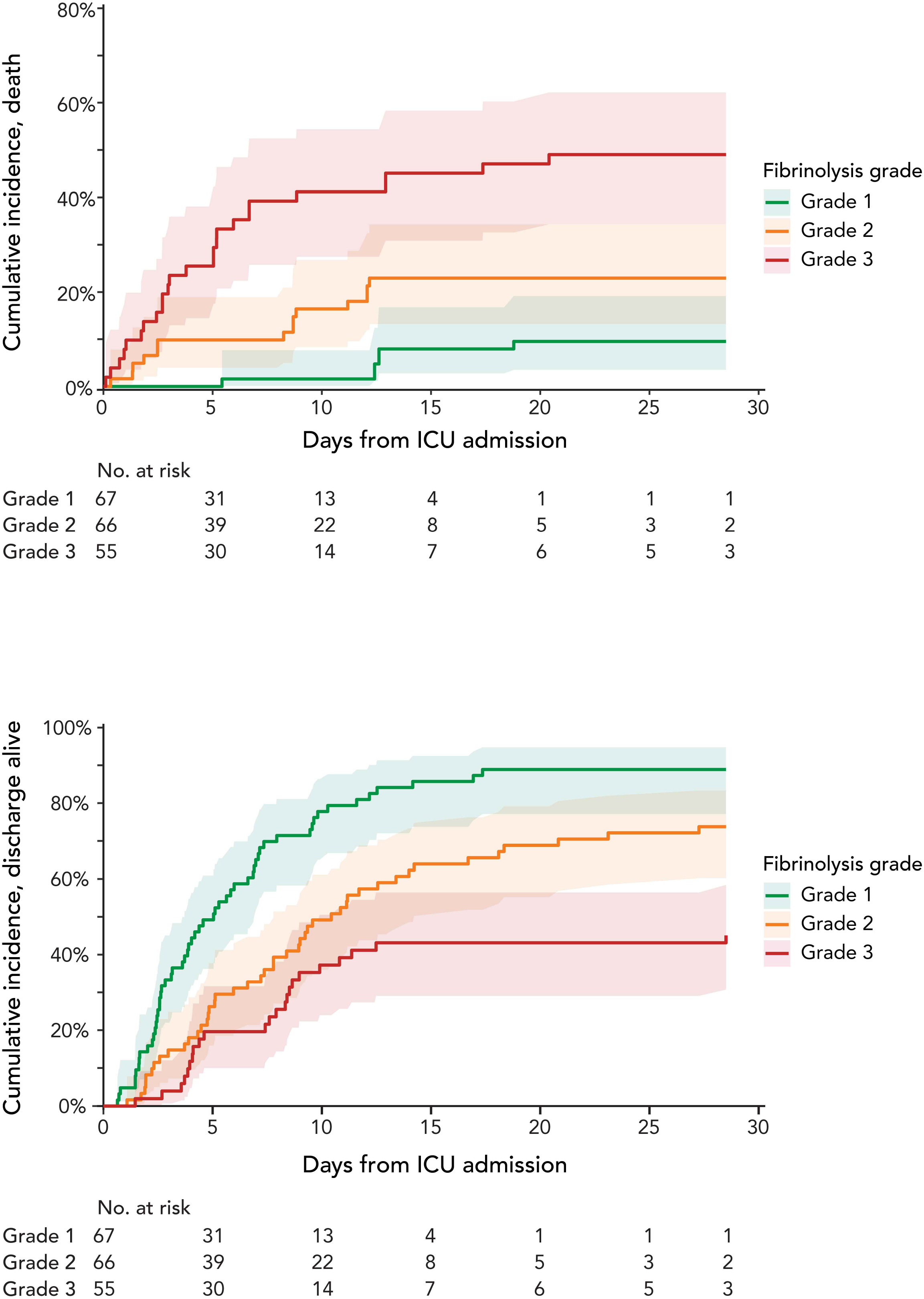
Cumulative incidence for death (top) and discharge alive (bottom) treated as competing events in the time-varying hazard ratio Cox regression using the fully adjusted (age, sex, anticoagulation, SOFA, time since admission to ICU for VET and steroids) model. Shaded areas indicate the 95% confidence interval. Green colour is used for Grade 1, orange for Grade 2 and red for Grade 3 fibrinolysis resistance. Patients at risk at each time point and for each Grade are indicated in the columns below each graph.

### Plasma fibrinolysis proteins

Plasma fibrinolytic regulators were measured in 283 samples from 69 patients across all grades of fibrinolysis resistance. Grade 3 fibrinolysis resistance was characterised by the greatest PAI-1/tPA and α2AP/plasminogen ratios with, respectively, 3.2 (2.1-4.9)-fold (p<0.001) and 1.8 (1.5-2.2)-fold (p<0.001) increases compared to Grade 1. Grade 2 fibrinolysis resistance demonstrated an intermediate state that remained significantly different from Grade 3 (see Additional File 1, Figure S3). These anti-fibrinolytic ratios provided the rationale for the specific *ex vivo* interventions to restore fibrinolytic capacity.

### Reversibility of fibrinolysis resistance ex vivo and translation to outcomes

The *ex vivo* effects of supplemental tPA, plasminogen or the α2AP iAb on fibrinolysis resistance were analysed in 36, 37 and 21 patients, respectively, and are shown in Figure 4 with details reported in the Additional File 1, Table S4. Additional tPA dosing (total dose 1.3 µg/ml), supplementing plasminogen or adding an α2AP iAb reduced fibrinolysis resistance in Grade 2 to Grade 1. In contrast, increased tPA dose was ineffective at improving fibrinolysis resistance in more than half of Grade 3 patients, whereas addition of plasminogen or α2AP iAb was effective in all Grade 3 patients.

**Figure 4.**
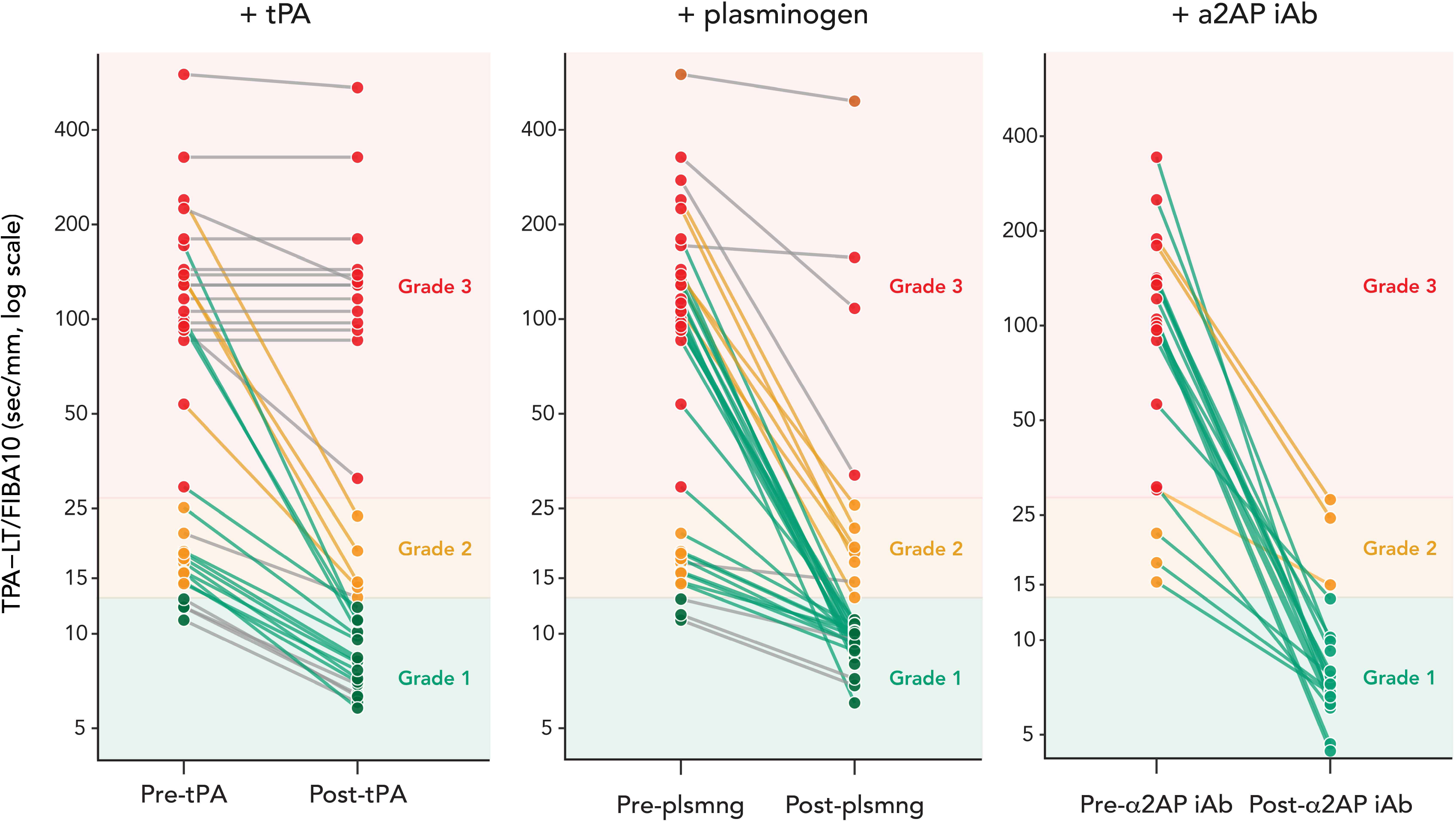
Change in the fibrin-adjusted lysis time (TPA-LT/FIBA10) before and after supplementation *ex vivo* with tPA (650 ng/mL), plasminogen (143 µg/mL) and an α2AP inhibitory antibody (iAb) (124 µg/mL, 5 µmol/L). Green colour indicates Grade 1, orange colour indicates Grade 2 and red colour indicates Grade 3 fibrinolysis resistance. Green connecting lines indicate an improvement in Grade of fibrinolysis resistance whereas grey connecting lines indicate lack of improvement across Grades.

A hypothetical interventional distribution framework was used to model changes in probability of death by translating the *ex vivo* effects of additional tPA, plasminogen or the α2AP iAb on the association between fibrinolysis resistance and mortality established in the observational cohort. The effects based on the continuous log(TPA-LT/FIBA10) measure are shown in Figure 5 and using the fully adjusted competing-risk model in Figure 6. Translated absolute and relative risk reductions for death and the numbers needed to treat from modelling the three *ex vivo* interventions are reported in Table 3. Additional tPA reduced the modelled predicted risk of death by 4.7 (3.7-14)% across Grades 2 and 3, driven by the 48% of assays that responded *ex vivo*; in Grade 3 the predicted probability of death was otherwise little changed, as no *ex vivo* effect was seen in the remaining 52% of assays with a non-significant effect overall. Supplemental plasminogen reduced the modelled predicted risk of death by 18 (8.1-24)% across Grades 2 and 3 and by 32 (15-42)% in Grade 3. The addition of α2AP iAb produced the greatest reduction in the modelled predicted risk of death by 21 (11-30)% across Grades 2 and 3 and by 36 (17-50)% in Grade 3. A full range of *ex vivo* effects translated by the model into the predicted risk of death can be explored at https://fortitude.shinyapps.io/FORTITUDESHINY/ (instructions in Additional File 1, Figure S4).

**Figure 5.**
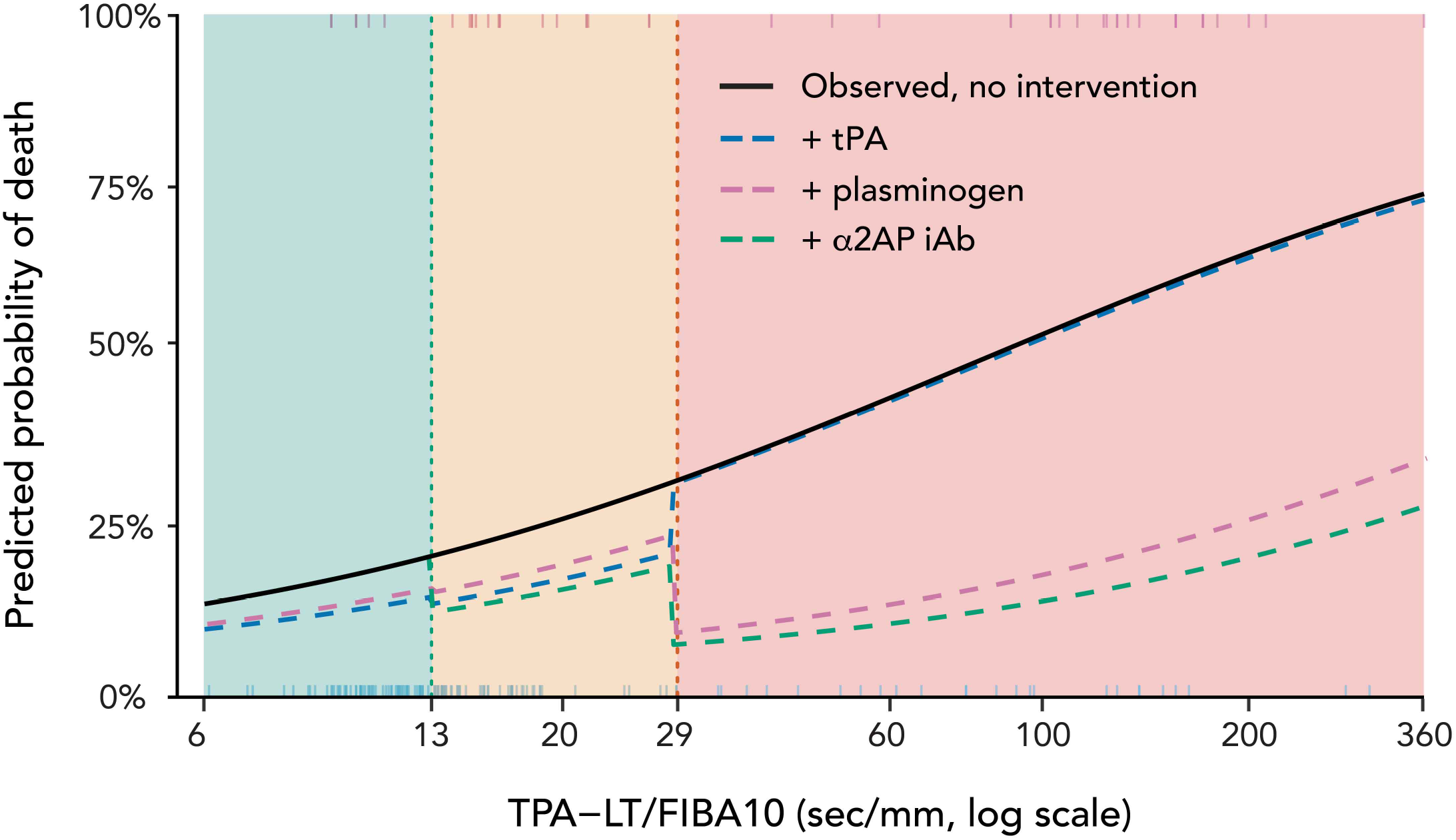
Changes in the predicted mortality up to 28 days associated with changes in the fibrinolysis resistance (continuous TPA-LT/FIBA10 on the log scale) modelled from the *ex vivo* effects (see Figure 4) of supplementation with additional tPA (blue), plasminogen (pink) or the inhibitory α2AP antibody (α2AP iAb, green) using a hypothetical intervention distribution framework (see also Figure 1). The solid black line represents the observed predicted mortality in the cohort without modelling any intervention effect. The dashed vertical lines represent the thresholds between the ordinal Grade 1 (green), Grade 2 (yellow) and Grade 3 (red) fibrinolysis resistance states. Note: no Grade 1 samples were tested for the α2AP iAb.

**Figure 6.**
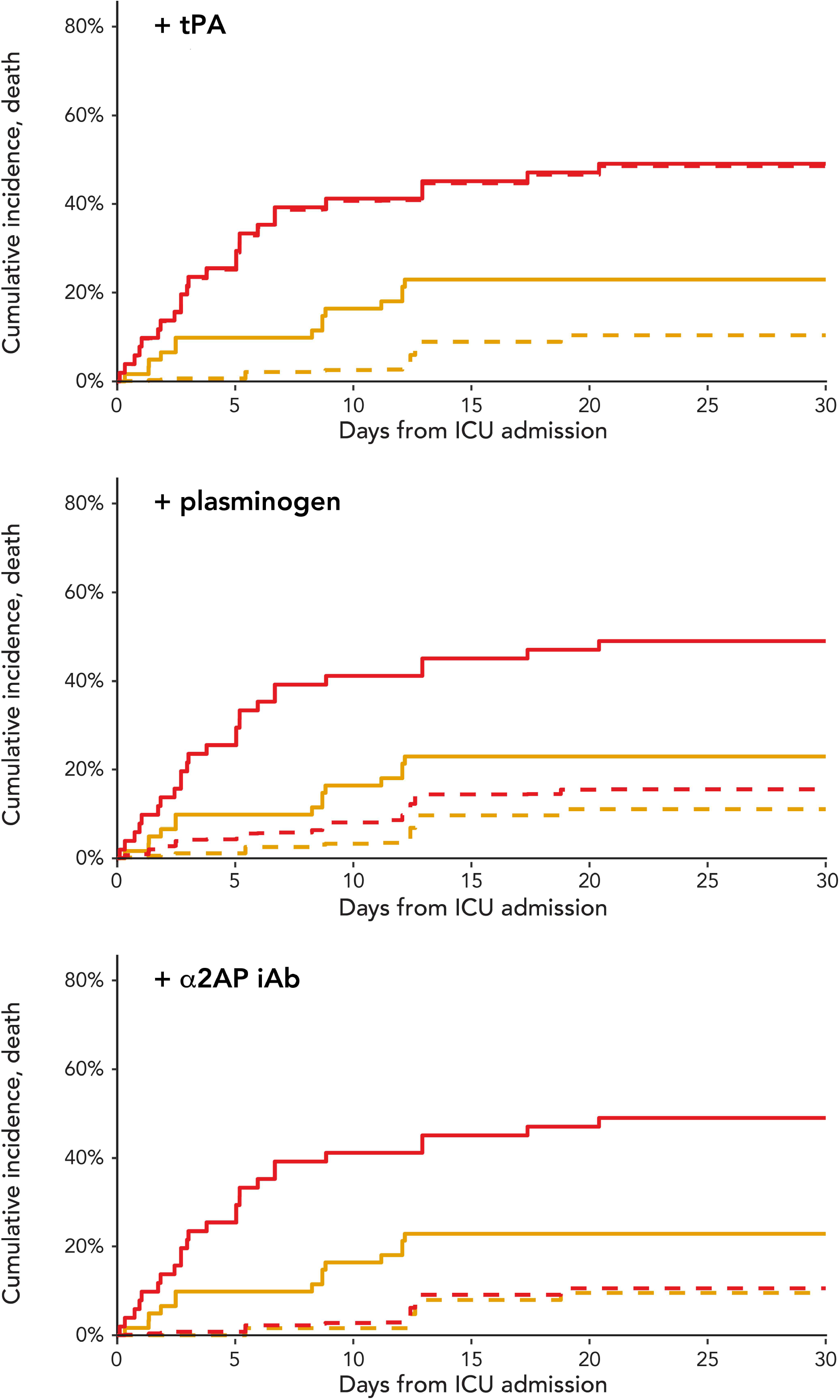
Changes in the cumulative incidence of death up to 28 days using a time-varying hazard ratio Cox regression model fully adjusted for age, sex, anticoagulation, SOFA, time since admission to ICU for VET and steroids, with discharge alive treated as a competing event. Changes were modelled from the *ex vivo* effects (see Figure 4) of supplementation with additional tPA, plasminogen or an inhibitory α2AP antibody (α2AP iAb). Red colour indicates Grade 3 fibrinolysis resistance and orange colour indicates Grade 2 fibrinolysis resistance. Solid lines represent the observed cumulative incidence without modelling any intervention effect, and dashed lines represent the effect using a hypothetical intervention distribution framework (see also Figure 1).

**Table 3.** Hypothetical intervention effect estimates by restoring fibrinolytic capacity. Effect estimates were obtained by modelling the *ex vivo* post-/pre-supplementation TPA-LT/FIBA10 ratios onto the fully adjusted competing-risks fibrinolysis resistance grade vs mortality association. The 95% CIs were generated from 1000 bootstrap iterations of both the *ex vivo* effect and the implied change in the fibrinolysis resistance to mortality association to account for uncertainty in both estimates. Baseline risk of death is the pre-intervention cumulative incidence for the fibrinolysis resistance grades.

| Intervention | Fibrinolysis resistance | Baseline risk of death, % (CIF<sup>a</sup>) | ARR<sup>b</sup>, % (95% CI) | RRR<sup>c</sup>, % | NNT<sup>d</sup> (95% CI) |
| --- | --- | --- | --- | --- | --- |
| + tPA | Grade 2+3 | 33 | 4.7 (3.7–14) | 11 | 27 (7.4–21) |
|  | Grade 3 | 49.5 | 5.3 (0.0–18.4) | NA | NA |
| + plasminogen | Grade 2+3 | 33 | 18 (8.1–24) | 53 | 5.7 (4.2–12) |
|  | Grade 3 | 49.5 | 32 (15–42) | 64 | 3.1 (2.4–6.7) |
| + α2AP iAb | Grade 2+3 | 33 | 21 (11–30) | 64 | 4.7 (3.3–8.8) |
|  | Grade 3 | 49.5 | 36 (17–50) | 73 | 2.8 (2.0–5.9) |

<sup>a</sup> CIF, cumulative incidence function; <sup>b</sup> ARR, absolute risk reduction; <sup>c</sup> RRR, relative risk reduction CI; <sup>d</sup> NNT, number needed to treat.

## DISCUSSION

This observational study of critically ill patients with sepsis has demonstrated that (1) increasing fibrinolysis resistance, quantified by the fibrin-adjusted tPA-enhanced VET lysis time and its derived three-tier grading system, was strongly and independently associated with 28-day mortality in a competing-risk model of the cohort; (2) the association followed a graded exposure-response relationship that persisted across unadjusted and adjusted models and remained robust to unmeasured confounding; (3) in a nested substudy, the most severe fibrinolysis resistance was substantiated by an imbalance of activators and inhibitors that favoured fibrinolysis inhibition with the highest ratio of PAI-1 to tPA activity and the highest ratio of α2AP to plasminogen concentrations; (4) restoring fibrinolysis balance *ex vivo* in the TPA-test using additional tPA alone had a limited effect confined to moderate (Grade 2) fibrinolysis resistance, whereas adding plasminogen or an α2AP inhibitory antibody (iAb) reversed fibrinolysis resistance even in the most severe (Grade 3) state; and (5) modelling these *ex vivo* effects onto the observational data within a hypothetical interventional distribution framework generated hypothetical reductions in 28-day mortality of 19–21% across Grades 2 and 3, and 32–36% in Grade 3.

The pathophysiology of sepsis-induced coagulopathy is increasingly understood to involve suppressed fibrinolysis, not only accelerated coagulation, as an important driver of microvascular thrombosis and organ failure [12]. Sepsis-induced coagulopathy is present in approximately one quarter of patients with sepsis and two-thirds of those with septic shock and is associated with poor outcomes and high mortality [1]. However, a specific measure of fibrinolysis resistance is lacking in current scoring systems for sepsis-induced and disseminated intravascular coagulopathy [17, 28], and these scores did not reliably distinguish patients across the grades of fibrinolysis resistance in this study. Fibrinolysis resistance in sepsis results from increased production of the fibrinolysis inhibitors PAI-1 and α2-antiplasmin [29–32] as well as decreased plasminogen levels from consumption [33, 34] and degradation by constituents of neutrophil extracellular traps [35]. The grading system of fibrinolysis resistance used in this study was closely aligned with these molecular signatures, supporting its use to rapidly ascertain the overall fibrinolysis state in whole blood within the critical care environment [6].

Importantly, fibrinolysis resistance determined by the point of care viscoelastometric assay was consistently aligned with the observed mortality in unadjusted and adjusted models. Thus, the results obtained in this cohort of sepsis patients, almost twice the size of the previous report of observational data only [6], further corroborates the dynamic associations with patient outcomes. The calculation of E-values addresses a key vulnerability of observational studies, being confounding by unmeasured variables that correlate with both the exposure and the outcome and hence could explain the observed associations [23]. The E-values for the risk ratios were robust at >3 for log(TPA-LT/FIBA10) and very strongly robust at 9-13 for Grade 3 vs Grade 1 fibrinolysis resistance [36, 37]. Any unmeasured confounder consistently associated with both fibrinolysis resistance and mortality would therefore need to generate a risk ratio >3 in a model already adjusted for age, sex, anticoagulation, SOFA score, VET timing and the administration of steroids. Confounding of this magnitude would need to represent a fundamentally different biological pathway in sepsis. Furthermore, the E-value >9 for the risk ratio comparing Grade 3 to Grade 1 patients, strongly suggests that the bedside grading tool identifies the primary endotype most likely associated with mortality and the one to target in any therapeutic efforts. Both plasminogen supplementation and administration of the inhibitory α2AP antibody to the TPA-test reduced fibrinolysis resistance to Grade 1 *ex vivo,* suggesting the potential of these interventions to be considered for clinical trials. In contrast, the potential therapeutic benefit in Grade 2 fibrinolysis resistance should be cautiously interpreted as the risk ratio in the fully adjusted time-varying model lost statistical significance. This is consistent with Grade 2 fibrinolysis resistance partially reflecting severity rather than a mechanistically distinct fibrinolytic endotype.

Therapeutic approaches should aim to correct substrate deficiency or relieve excessive inhibition of plasmin generation rather than directly driving thrombolysis, thereby rebalancing fibrinolytic activity with potentially less systemic lysis and bleeding risk. The *ex vivo* experiments illustrate how the principal mechanisms of fibrinolysis resistance in sepsis may be targeted. A moderately responsive endotype to double tPA dosing was restricted to Grade 2. The addition of plasminogen restored fibrinolysis capacity in both Grade 2 and even the most severe Grade 3 fibrinolysis resistance states. This effect was even more pronounced using an α2AP inhibitory antibody. This supports plasminogen supplementation or α2AP inhibition, rather than increased tPA, as the more rational interventional strategies in both Grade 2 and Grade 3 resistance. Very high PAI-1 levels in some Grade 2 cases might limit the ability to restore fibrinolysis through administration of tPA alone without increasing the risk of bleeding. Vanguard studies on profibrinolytic therapy in critically ill patients with acute infection [13, 38, 39] have not been broadly adopted due to a perceived bleeding risk, although no clinically significant adverse bleeding episodes were reported. The point-of-care VET used in this study enables rapid detection, ongoing monitoring and grading of fibrinolysis resistance in addition to whole blood coagulation and, therefore, may help alleviate concerns for bleeding complications through precision guided patient selection and dosing.

Conventional observational analyses can estimate associations between fibrinolysis resistance and mortality, but do not address what might happen if fibrinolysis was changed by an intervention. The hypothetical interventional distribution framework was applied in this study as a principled method to fill this gap by a counterfactual analysis. Each patient’s fibrinolysis resistance was shifted according to the magnitude of effect observed in the *ex vivo* experiments, and the mortality predicted from the observational exposure-response model updated to provide quantitative, competing-risks-based estimates. This makes no causal claim beyond what the data can support, but allows the *ex vivo* and observational findings to be expressed on a single, clinically interpretable scale [25, 26].

By modelling the *ex vivo* effects of plasminogen or an α2AP iAb onto the observational data, we estimated hypothetical reductions in 28-day mortality corresponding to numbers needed to treat of 5-6 across Grades 2 and 3 fibrinolysis resistance, and 3 in Grade 3 resistance. A recent pilot study of plasminogen-containing plasma supplementation in sepsis significantly raised functional plasminogen and plasmin generation without changing PAI-1 or tPA levels [11], congruent with the substrate-repletion mechanism implied in the *ex vivo* plasminogen effect in this study. Notably, that trial reported a non-significant mortality signal favouring plasminogen supplementation (42.3% vs 60.0%; the number needed to treat being 6) that is directionally concordant with the absolute risk reductions estimated in the present hypothetical intervention framework. The time-varying Cox hazard ratio model used in this study support early interventions, preferably within 72 hours of admission to ICU given the steep increase in mortality during this period. This therapeutic window is highly feasible in a clinical context in which tPA-enhanced VET is employed and would allow for patient identification, risk stratification and commencing targeted therapies to restore fibrinolysis balance.

### Implications for a future clinical trial

The interventions used in this study may help model pro-fibrinolytic interventions in a clinical trial, with both plasminogen (Ryplazim®, Kedrion Biopharma, Italy) and α2AP iAb (NCT06149520),[40] available for clinical use. A future trial should target patients with Grade 3 fibrinolysis resistance given that this endotype had the most robust association with 28-day mortality. The steep increase in the cumulative incidence of death within 5 days supports early intervention. The modelled hypothetical-intervention effects represent the maximal benefit assuming complete translation of *ex vivo* reversal to clinical outcome. This is biologically unlikely, and a more plausible effect size is a 10% absolute risk reduction from the observed control mortality of 46%. In a two-arm, 1:1 placebo-controlled superiority trial with 90% power and a two-sided α of 0.05, 507 patients per arm would be required (1014 total; ∼1116 allowing 10% attrition). The observed prevalence of Grade 3 fibrinolysis resistance at 31% means about 3600 patients would need to be screened that is feasible within a multicentre study design. A range of effect sizes is shown in the Additional File 1, Figure S5.

### Strengths and limitations

This study extends the recently described use of a tPA enhanced VET assay for the development of a grading system for fibrinolysis resistance [6], to identify potential interventions in targeted patients. Unadjusted and adjusted competing risk models of time-varying hazard ratio for death within 28 days identified an early therapeutic window to target fibrinolysis resistance after admission to ICU for sepsis. Computed E-values were robust to plausible unmeasured confounding for the associations with mortality using the continuous log(TPA-LT/FIBA10) and the Grade 3 versus Grade 1 comparison. Because the study included time-varying confounders and competing risks, these values should still be interpreted cautiously as approximate measures of robustness. The results of this study must be viewed as hypothesis-generating rather than predictive of clinical trial outcomes. The hypothetical interventional distribution framework rests on the assumption that the observed association between fibrinolysis resistance and death is at least partly causal and that pharmacological reversal of the experimental endotype would translate into a commensurate reduction in mortality. This assumption cannot be tested with observational data, however granular. The *ex vivo* effects were measured in whole-blood assays with reagents optimised to evaluate coagulation and fibrinolysis within a point-of-care time frame and, hence, do not necessarily represent physiological concentrations or reaction kinetics occurring *in vivo*. Only fixed doses of pro-fibrinolytic agents were examined. The immediate biochemical reversibility of fibrinolysis is captured in the *ex vivo* assays, while the downstream *in vivo* consequences of restored lysis, that could include lysis of haemostatic clots and bleeding, are not. The impacts and actions of non-soluble endothelial, vascular factors are excluded. Furthermore, the viscoelastometric assays examine the lysis of the clot as it is formed and might not be representative of the clinical situation in which mature clots are present. The hypothetical intervention framework does not model the competing risk of bleeding which is a central safety consideration for any fibrinolytic strategy in critically ill patients. Finally, plasmin activity is differentially regulated during the survival and repair stages of the acute phase response and promoting increased generation of plasmin early might disrupt the tissue damage matrix that provides haemostasis, containment of microorganisms [41] and a scaffold for subsequent tissue repair [42].

This was a single-centre study, and the fibrinolysis resistance thresholds require external validation. The sample size yielded wide confidence intervals for some time-specific effect estimates on mortality, and longer-term and functional outcomes were not examined.

## CONCLUSIONS

This study demonstrated that fibrinolysis resistance measured at the bedside is associated with 28-day mortality in critically ill patients with sepsis and is reversible *ex vivo*. Embedding experimental reversibility within a competing-risks model based on a clinical cohort of 188 patients enabled modelling of quantitative, confounding-aware estimates of the potential mortality benefit of restoring fibrinolytic capacity. These estimates are conditional and hypothesis-generating but identify plasminogen supplementation or α2AP inhibition as candidate interventions. Taken together, these findings support the design of a clinical trial that aligns interventions with biological pathways, considers dynamic pathophysiological trajectories, and allows for enrichment strategies based on responsive endotypes with a clinically meaningful and plausible survival benefit.

## LIST OF ABBREVIATIONS

α2AP: iAb Inhibitory antibody to alpha-2-antiplasmin
FIBA10: Fibrin clot amplitude at 10 minutes
PAI-1: Plasminogen activator inhibitor type-1
tPA: Tissue plasminogen activator
TPA-LT: Lysis time in the TPA-test of fibrinolysis
VET: Viscoelastometric test

## DECLARATIONS

### Ethics approval and consent to participate

The study was approved by the Human Ethics Research Committee, South Western Sydney Local Health District (2022/ETH02122) with waiver of participants’ informed consent as permitted under the National Statement on Ethical Conduct in Human Research 2023 for low-risk research in critically ill patients. The research program is registered on Clinicaltrials.gov (NCT06680180).

### Consent for publication

Not applicable.

### Availability of data and materials

The datasets used are available from the corresponding author on reasonable request and as permitted by the local Ethics Governance Board. A synthetic dataset generated from the actual data that can be used together with the hypothetical intervention distribution framework RStudio code is available at: https://zenodo.org/records/22821116. An interactive analysis of the data is available at: https://fortitude.shinyapps.io/FORTITUDESHINY/

### Authors contributions

LC: Project conception, design, funding, conduct, data collection, analyses and manuscript preparation. MS: Patient screening, data collection and manuscript preparation. ES: Patients, screening, data collection and database management. RL: Intellectual input and manuscript preparation. PC: Intellectual input and manuscript preparation. CK: Data analyses and ELISA assay supervision, intellectual input and manuscript preparation. SF: Data analyses and manuscript preparation. DR: Intellectual input and manuscript preparation. PKM: Intellectual input and manuscript preparation. RM: Data analyses and ELISA assay supervision, intellectual input and manuscript preparation. AA: Project conception, design, conduct, clinical supervision, data collection, analyses and manuscript preparation. All authors read and approved the final manuscript.

## Acknowledgements

The authors extend their sincere gratitude to the patients and their families who participated in this study. We are grateful to the nursing and medical staff of the Liverpool Hospital Intensive Care Unit for their assistance with this study, in particular the ClotPro Champions, Intensive Care nurses who were trained on the ClotPro technology and provided bedside capability: Alexandra Heath, Emma Hill, Marcus Iedani, Karla Lopez, Elisa McClure, Jennifer McClure, Yvette McManus, Carlos Mendoza, Jennene Miller, Mostafa Hasha, Sharyn Parker, Julio Ruiz Davila, Krystle-Lee Strickland, Anne Tran, Bija Unnikrishnan, Harley Whitfield, Sanjay Singh, Katie Mills. Joanne Chia, Tammy Lam, Zikou Liu for performing the ELISA assays. Mrs Korina Valentine, consumer representative, provided essential input to the project design as a survivor of sepsis.

## Additional file 1

**Table S1.**
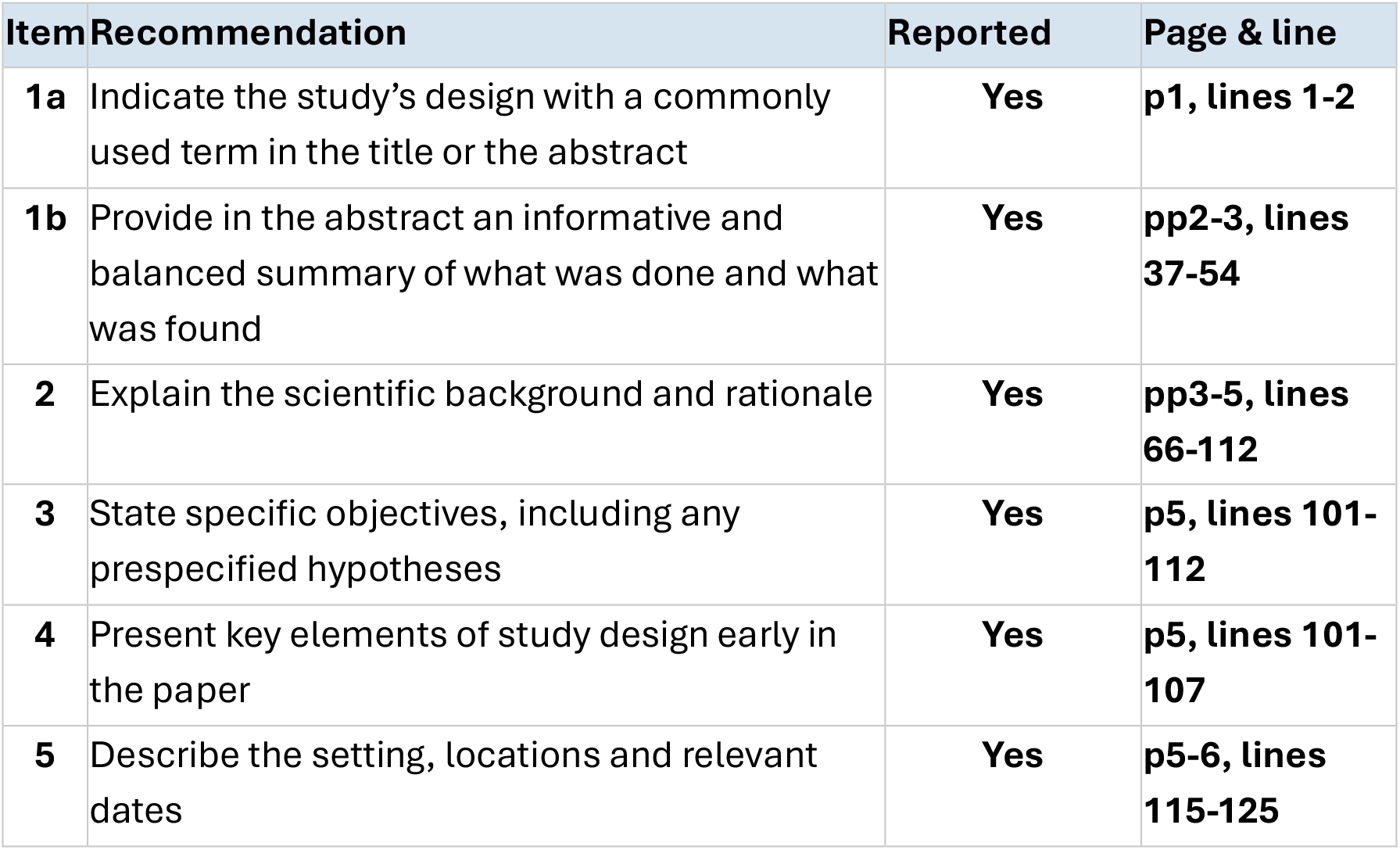
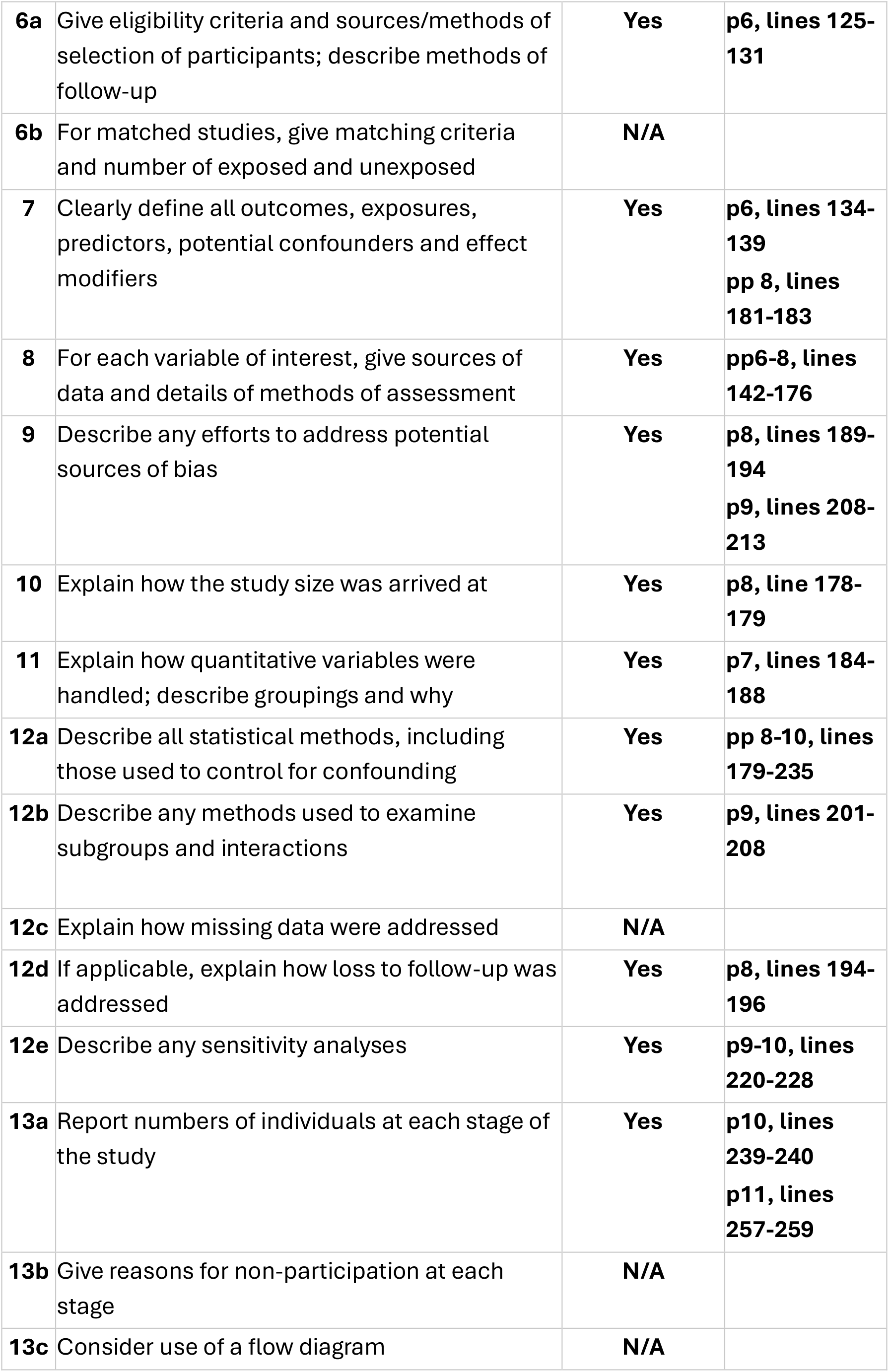
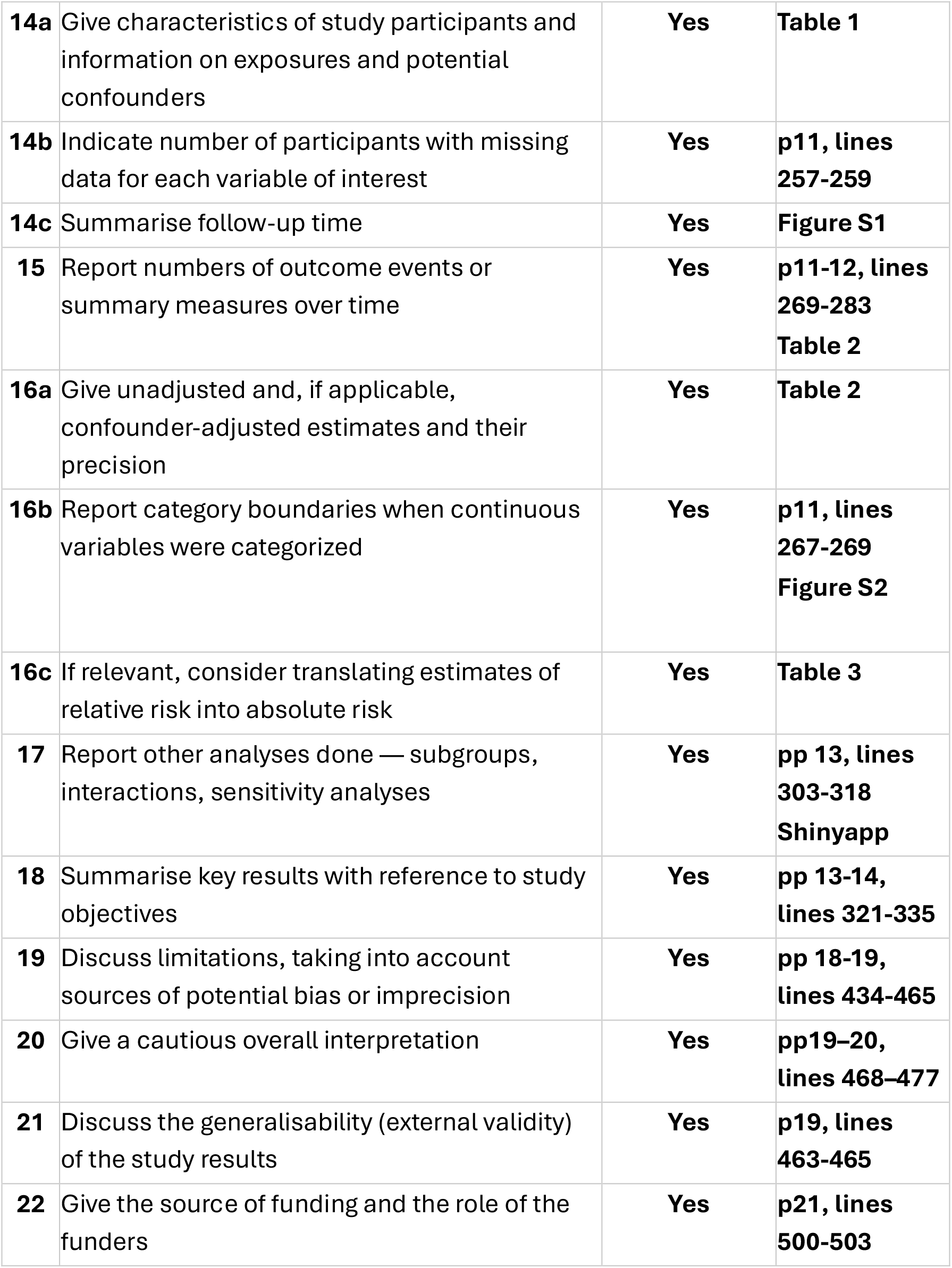
STROBE checklist. Page and line numbers refer to the submitted Word document containing the text body of the manuscript.

| Item | Recommendation | Reported | Page & line |
| --- | --- | --- | --- |
| **1a** | Indicate the study's design with a commonly used term in the title or the abstract | **Yes** | **p1, lines 1-2** |
| **1b** | Provide in the abstract an informative and balanced summary of what was done and what was found | **Yes** | **pp2-3, lines 37-54** |
| **2** | Explain the scientific background and rationale | **Yes** | **pp3-5, lines 66-112** |
| **3** | State specific objectives, including any prespecified hypotheses | **Yes** | **p5, lines 101-112** |
| **4** | Present key elements of study design early in the paper | **Yes** | **p5, lines 101-107** |
| **5** | Describe the setting, locations and relevant dates | **Yes** | **p5-6, lines 115-125** |
| **6a** | Give eligibility criteria and sources/methods of selection of participants; describe methods of follow-up | **Yes** | **p6, lines 125-131** |
| **6b** | For matched studies, give matching criteria and number of exposed and unexposed | **N/A** |  |
| **7** | Clearly define all outcomes, exposures, predictors, potential confounders and effect modifiers | **Yes** | **p6, lines 134-139**<br>**pp 8, lines 181-183** |
| **8** | For each variable of interest, give sources of data and details of methods of assessment | **Yes** | **pp6-8, lines 142-176** |
| **9** | Describe any efforts to address potential sources of bias | **Yes** | **p8, lines 189-194**<br>**p9, lines 208-213** |
| **10** | Explain how the study size was arrived at | **Yes** | **p8, line 178-179** |
| **11** | Explain how quantitative variables were handled; describe groupings and why | **Yes** | **p7, lines 184-188** |
| **12a** | Describe all statistical methods, including those used to control for confounding | **Yes** | **pp 8-10, lines 179-235** |
| **12b** | Describe any methods used to examine subgroups and interactions | **Yes** | **p9, lines 201-208** |
| **12c** | Explain how missing data were addressed | **N/A** |  |
| **12d** | If applicable, explain how loss to follow-up was addressed | **Yes** | **p8, lines 194-196** |
| **12e** | Describe any sensitivity analyses | **Yes** | **p9-10, lines 220-228** |
| **13a** | Report numbers of individuals at each stage of the study | **Yes** | **p10, lines 239-240**<br>**p11, lines 257-259** |
| **13b** | Give reasons for non-participation at each stage | **N/A** |  |
| **13c** | Consider use of a flow diagram | **N/A** |  |
| **14a** | Give characteristics of study participants and information on exposures and potential confounders | **Yes** | **Table 1** |
| **14b** | Indicate number of participants with missing data for each variable of interest | **Yes** | **p11, lines 257-259** |
| **14c** | Summarise follow-up time | **Yes** | **Figure S1** |
| **15** | Report numbers of outcome events or summary measures over time | **Yes** | **p11-12, lines 269-283**<br>**Table 2** |
| **16a** | Give unadjusted and, if applicable, confounder-adjusted estimates and their precision | **Yes** | **Table 2** |
| **16b** | Report category boundaries when continuous variables were categorized | **Yes** | **p11, lines 267-269**<br>**Figure S2** |
| **16c** | If relevant, consider translating estimates of relative risk into absolute risk | **Yes** | **Table 3** |
| **17** | Report other analyses done — subgroups, interactions, sensitivity analyses | **Yes** | **pp 13, lines 303-318**<br>**Shinyapp** |
| **18** | Summarise key results with reference to study objectives | **Yes** | **pp 13-14, lines 321-335** |
| **19** | Discuss limitations, taking into account sources of potential bias or imprecision | **Yes** | **pp 18-19, lines 434-465** |
| **20** | Give a cautious overall interpretation | **Yes** | **pp19–20, lines 468–477** |
| **21** | Discuss the generalisability (external validity) of the study results | **Yes** | **p19, lines 463-465** |
| **22** | Give the source of funding and the role of the funders | **Yes** | **p21, lines 500-503** |

**Table S2.**
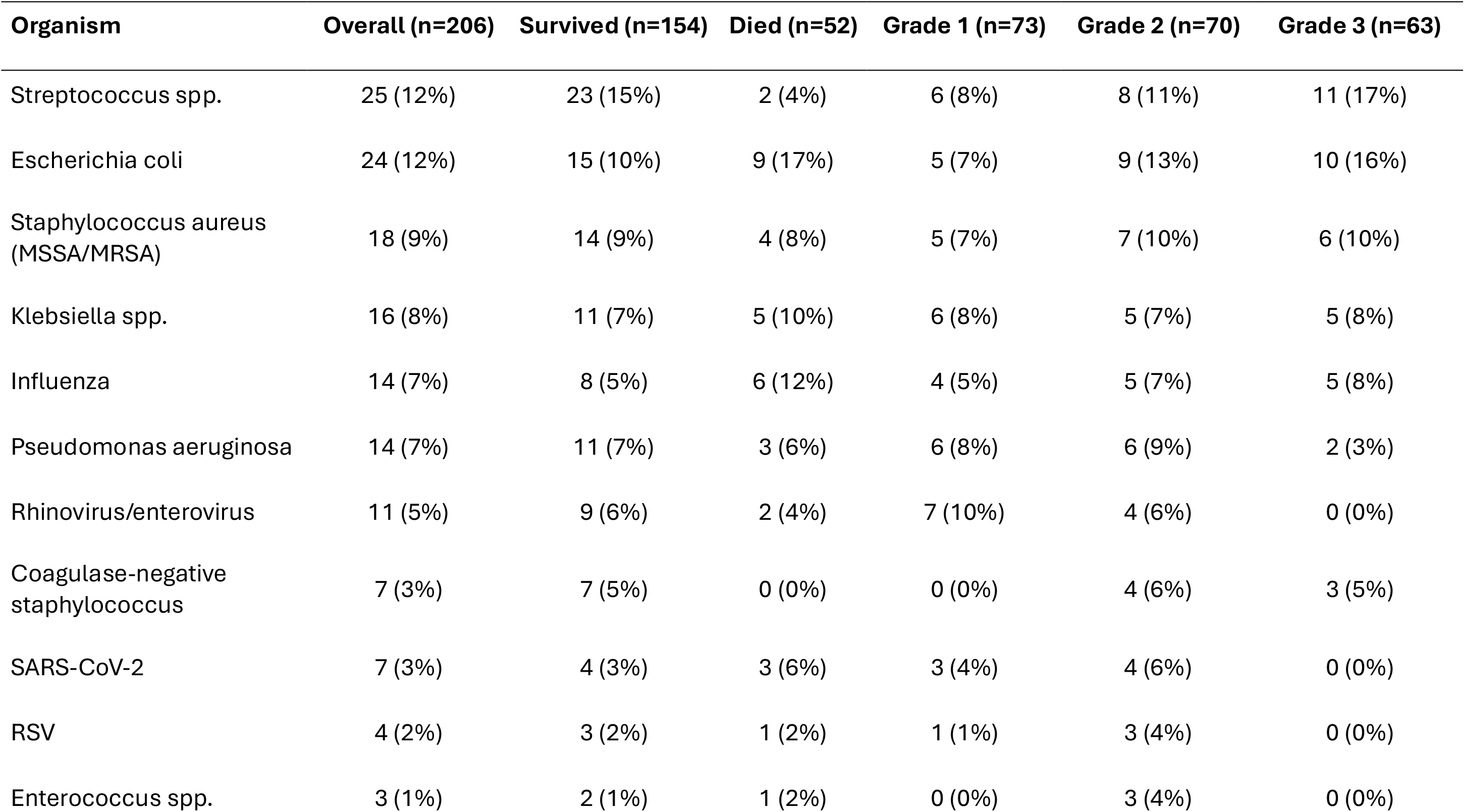
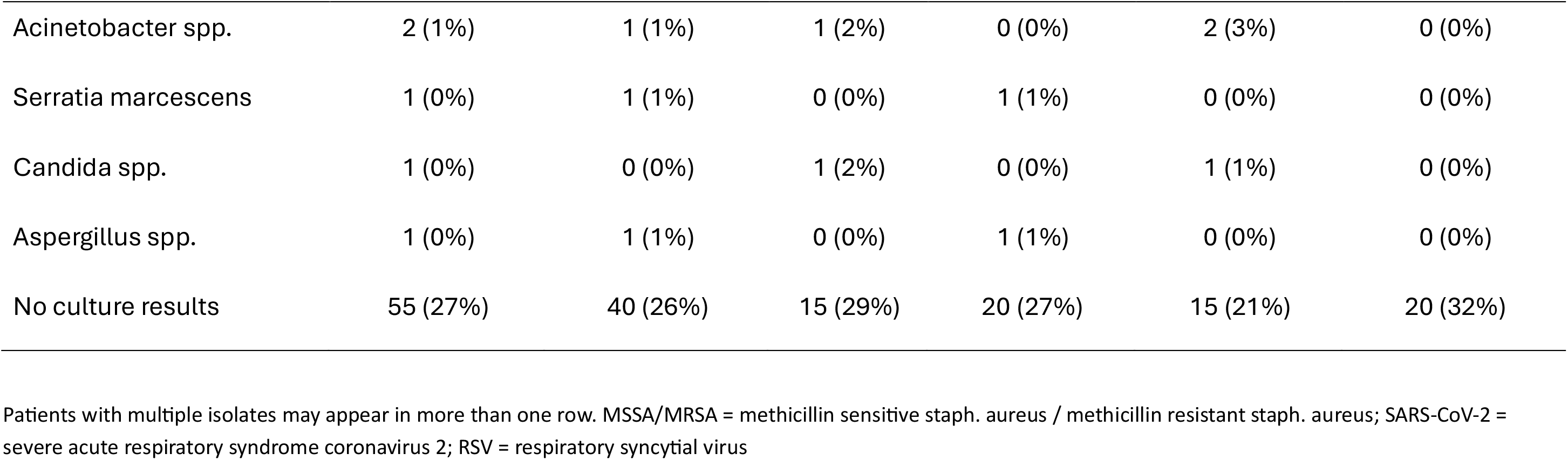
Microbial isolates. Data are reported overall and split by survival and fibrinolysis resistance grades.

**Table S3.** Characteristics of excluded patients. The 18 patients excluded from the adjusted models of the association between fibrinolysis resistance and death up to 28 days were not significantly different from the complete, observational cohort (n=188). Data are median and interquartile range.

| **Variable** | **Retained** | **Excluded** | **p-value** |
| --- | --- | --- | --- |
| **n** | **n=188** | **n=18** |  |
| Admission APACHE III | 82 [66-100] | 84 [63-100] | 0.96 |
| Admission SOFA | 12 [9-14] | 14 [10-14] | 0.40 |
| First TPA-LT/FIBA10 (mm/sec) | 15 [11-41] | 20 [12-38] | 0.91 |
| Grade 3 at first VET, n (%) | 55 (29) | 8 (44) | 0.19 |
| Death up to 28 days, n (%) | 48 (26) | 4 (22) | 1.00 |

**Table S4.** *Ex vivo* reversibility of fibrinolysis resistance. The capacity of pro-fibrinolytic agents to reverse fibrinolysis resistance were tested *ex vivo* and are reported by intervention and split by fibrinolysis resistance grade. The number of patients are indicated by n= and values are reported as median [IQR]. The p-values next to the intervention labels report the within-intervention comparison by the Kruskal-Wallis test between ratio. The between-intervention comparison for the most severe (Grade 3) fibrinolysis resistance is reported by the Mann-Whitney test.

| Fibrinolysis resistance | n | Pre TPA-LT/FibA10 | Post TPA-LT/FibA10 | Ratio (post/pre) | Absolute reduction | Responders |
| --- | --- | --- | --- | --- | --- | --- |
| **+ tPA (n=36), p=0.37** |  |  |  |  |  |  |
| Grade 1 | 4 | 12.2 [11.9–12.3] | 6.38 [6.26–6.54] | 0.54 [0.52–0.55] | 5.5 [5.2–5.9] | 4/4 (100%) |
| Grade 2 | 11 | 17.3 [15.7–19.67] | 8.17 [7.42–9.70] | 0.47 [0.42–0.52] | 9.2 [7.7–9.7] | 11/11 (100%) |
| Grade 3 | 21 | 129 [97.3–171] | 97.3 [18.3–132] | 1.00 [0.14–1.00] | 0.0 [0.0–89.9] | 10/21 (48%) |
| **+ Plasminogen (n=37), p<0.001** |  |  |  |  |  |  |
| Grade 1 | 3 | 11.5 [11.3–12.2] | 7.19 [7.01–8.37] | 0.62 [0.62–0.68] | 4.2 [3.8–4.3] | 3/3 (100%) |
| Grade 2 | 10 | 17.1 [15.6–18.2] | 10.0 [9.43–10.3] | 0.58 [0.54–0.62] | 7.0 [5.7–8.0] | 10/10 (100%) |
| Grade 3 | 24 | 129 [99.3–174] | 10.8 [9.1–19.6] | 0.10 [0.07–0.14] | 107 [87–127] | 24/24 (100%) |
| Grade 3 (tPA) + plasminogen vs Grade 3 (tPA) + tPA, p<001 |  |  |  |  |  |  |
| **+ α2AP iAb (n=21), p=001** |  |  |  |  |  |  |

| **Fibrinolysis resistance** | **n** | **Pre TPA-LT/FibA10** | **Post TPA-LT/FibA10** | **Ratio (post/pre)** | **Absolute reduction** | **Responders** |
| --- | --- | --- | --- | --- | --- | --- |
| Grade 2 | 5 | 20.9 [16.8–28.7] | 6.60 [6.49–7.39] | 0.39 [0.35–0.44] | 11.7 [9.3–16.0] | 5/5 (100%) |
| Grade 3 | 16 | 122 [94.1–144] | 7.15 [6.20–9.54] | 0.06 [0.05–0.09] | 110 [86–135] | 16/16 (100%) |
| Grade 3 (tPA) + α2AP iAb vs (tPA) + tPA, p<0.001; vs (tPA) + plasminogen, p=0.006 | | | | | | |

**Figure S1.**
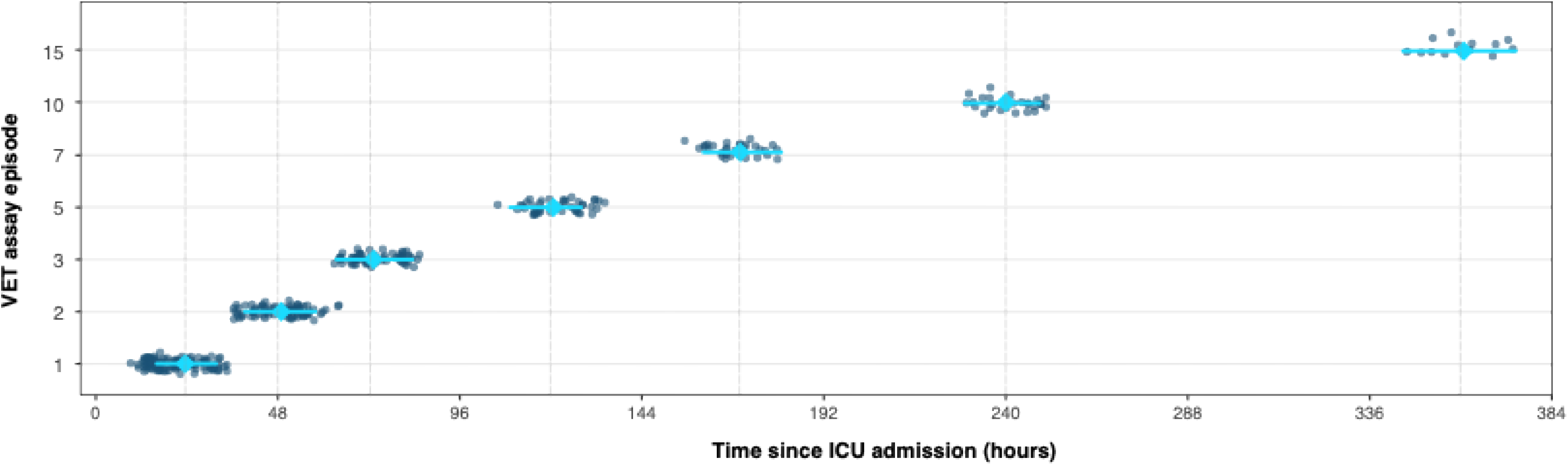
Timing of viscoelastometric assays.

**Figure S2.**
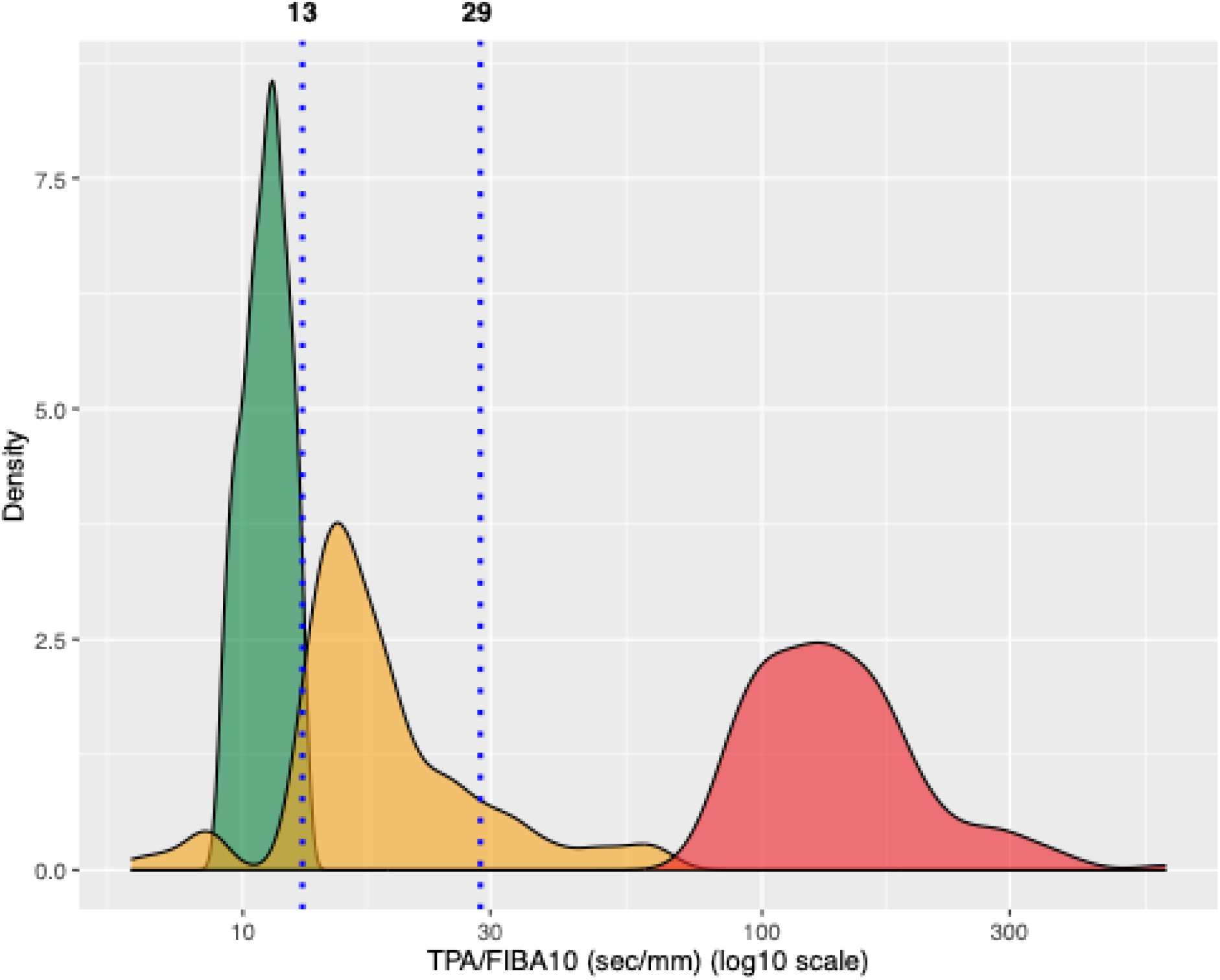
Cluster analysis. Fibrinolysis resistance was quantified by the ratio between log-transformed lysis time (TPA-LT, sec) and fibrinogen clot amplitude at 10 minutes (FIBA10). Data-driven, unsupervised univariate cluster analysis was performed using a Gaussian mixture model and Box-Cox transformed data. Grading of fibrinolysis resistance was performed according to the clusters as Grade 1 <13 sec/mm (green), Grade 2 ≥13 and <29 sec/mm (organge) and Grade 3 ≥29 (red). See also reference [5].

**Figure S3.**
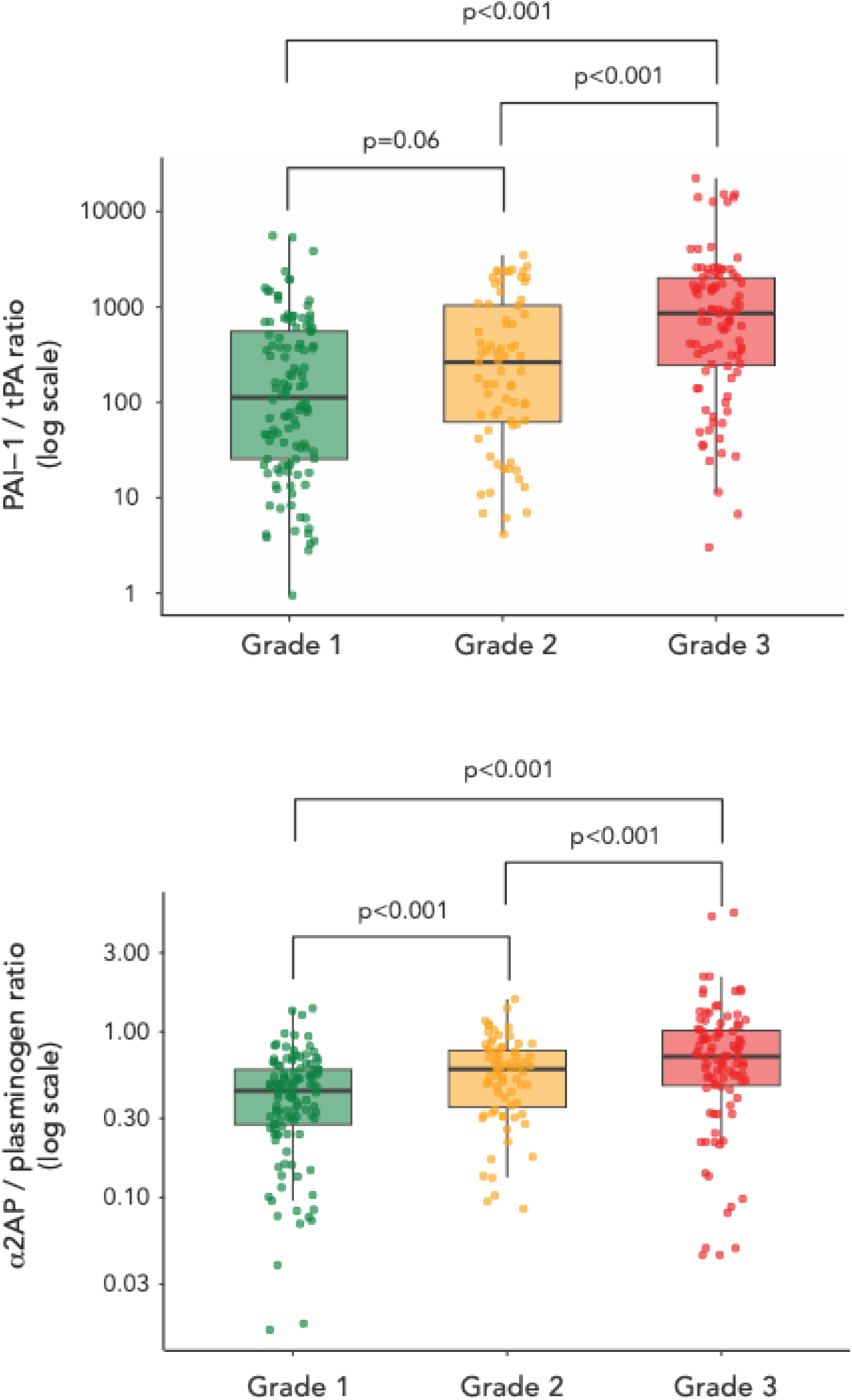
Key fibrinolysis regulator balance. Plasma levels of key fibrinolysis proteins were measured in 69 patients across the three grades (Grade 1, green; Grade 2, orange; Grade 3 red) of fibrinolysis resistance (263 assays in total). The PAI-1 to tPA ratio increased 1.45 (0.98-2.15)-fold in Grade 2 vs 1, and increased 3.17 92.07-4.85)-fold in Grade 3 vs 1. The α2AP (α2-antiplasmin) to plasminogen ratio increased 1.33 (1.14-1.54)-fold in Grade 2 vs 1, with a further increase to 1.79 (1.52-2.11)-fold in Grade 3 vs 1. Differences in fibrinolysis protein ratios were analysed by a linear mixed-effects model with a random intercept for patient to account for within-subject correlation.

**Figure S4.**
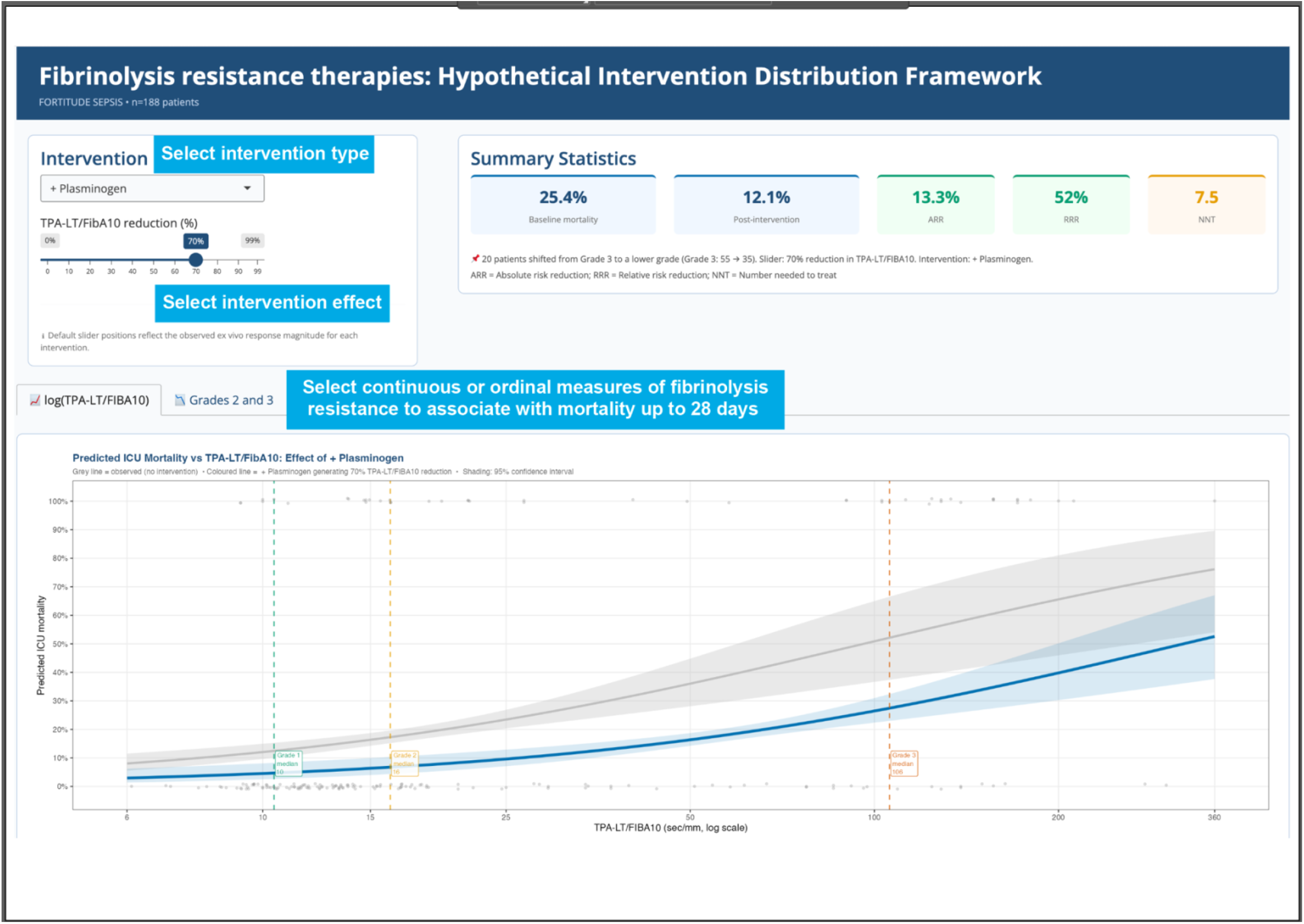
Interactive simulation of hypothetical intervention effects.

**Figure S5.**
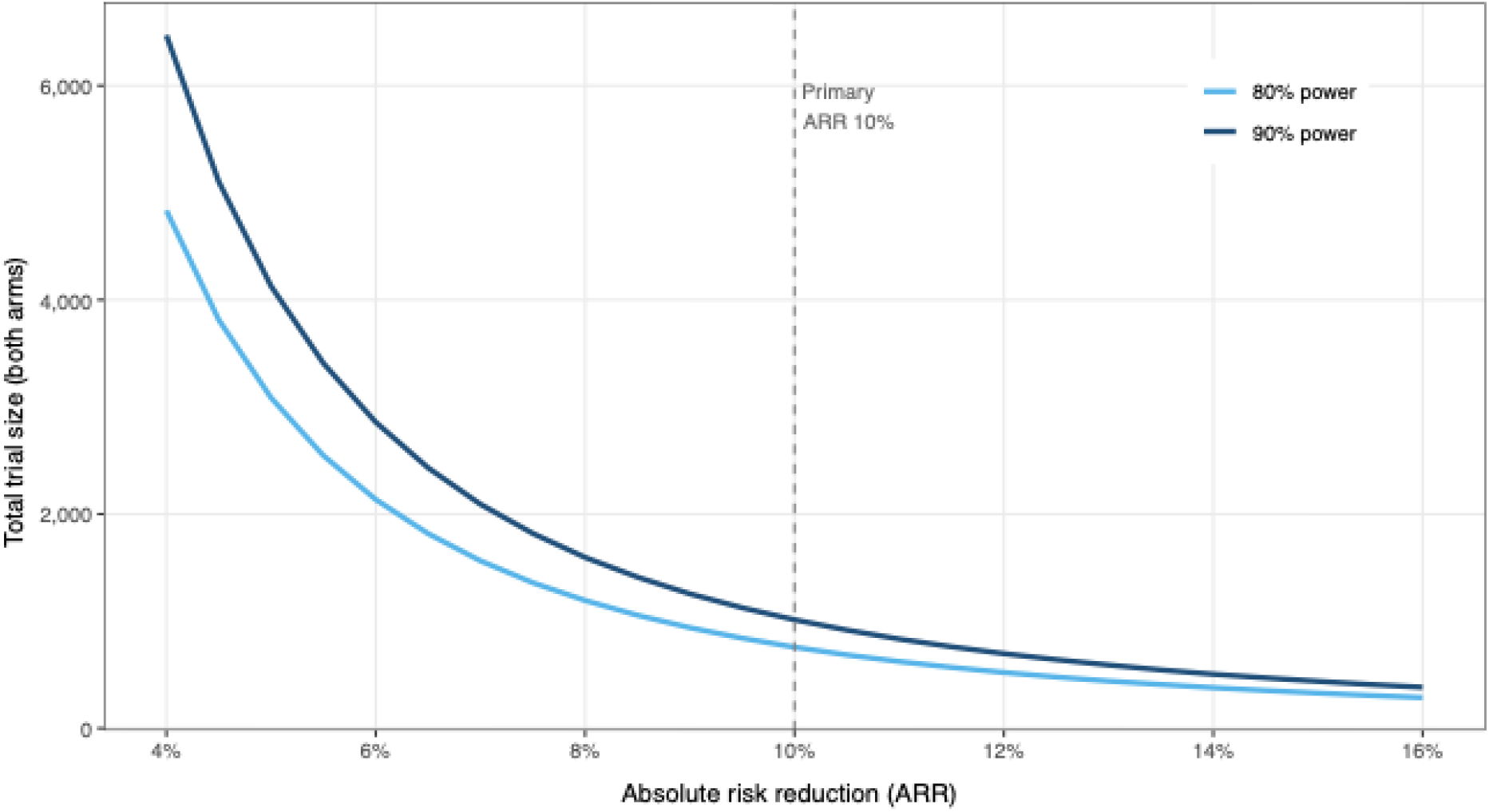
Trial size calculations. Trial sizes in a two-arm, 1:1 placebo-controlled superiority design at a=0.05 and power of 80% (light blue) or 90% (dark blue) for ranges of absolute risk reduction in mortality up o 28 days.

## REFERENCES

1. Williams B, Zou L, Pittet JF, Chao W, (2024) Sepsis-Induced Coagulopathy: A Comprehensive Narrative Review of Pathophysiology, Clinical Presentation, Diagnosis, and Management Strategies. Anesth Analg 138: 696–711

2. Moore HB, (2023) Fibrinolysis Shutdown and Hypofibrinolysis Are Not Synonymous Terms: The Clinical Significance of Differentiating Low Fibrinolytic States. Semin Thromb Hemost 49: 433–443

3. Iba T, Umemura Y, Wada H, Levy JH, (2021) Roles of Coagulation Abnormalities and Microthrombosis in Sepsis: Pathophysiology, Diagnosis, and Treatment. Arch Med Res 52: 788–797

4. Marshall JC, (2014) Why have clinical trials in sepsis failed? Trends Mol Med 20: 195–203

5. David S, Leone M, Girardis M, Muller MM, Gavrilovic S, Domizi R, Damiani E, Martin-Loeches I, Ferrer R, Bode C, Chousterman B, Russell L, (2026) Innovation in sepsis trials: rethinking endpoints, statistics and patient stratification. Intensive Care Med Exp 14

6. Coupland LA, Frost SA, Lin J, Pham N, Self M, Crispin PJ, Rabbolini DJ, Keragala CB, Medcalf RL, Aneman A, (2026) A grading system of dynamic fibrinolysis resistance in sepsis associates with ICU outcomes. Am J Respir Crit Care Med

7. Coupland LA, Rabbolini DJ, Schoenecker JG, Crispin PJ, Miller JJ, Ghent T, Medcalf RL, Aneman AE, (2023) Point-of-care diagnosis and monitoring of fibrinolysis resistance in the critically ill: results from a feasibility study. Crit Care 27: 55

8. Self M, Coupland LA, Aneman A, (2025) Utility of fibrinolysis enhanced viscoelastic assays to evaluate fibrinolysis disorders in critically ill adults with severe infection: a scoping review. Ann Intensive Care 15: 110

9. Schmitt FCF, Manolov V, Morgenstern J, Fleming T, Heitmeier S, Uhle F, Al-Saeedi M, Hackert T, Bruckner T, Schochl H, Weigand MA, Hofer S, Brenner T, (2019) Acute fibrinolysis shutdown occurs early in septic shock and is associated with increased morbidity and mortality: results of an observational pilot study. Ann Intensive Care 9: 19

10. Hvas CL, Larsen JB, (2023) The Fibrinolytic System and Its Measurement: History, Current Uses and Future Directions for Diagnosis and Treatment. Int J Mol Sci 24

11. Tschirhart M, Curtiaud A, Abou Rjeily R, Merdji H, Demiselle J, Severac F, Guerin D, Cacao M, El Ghazouani F, Angles-Cano E, Toti F, Meziani F, Helms J, (2026) Plasminogen supplementation reverses fibrinolytic insufficiency in sepsis-induced disseminated intravascular coagulation: a pilot study. Intensive Care Med 52: 456–465

12. Tschirhart M, Martin M, Curtiaud A, Angles-Cano E, Meziani F, Toti F, Helms J, (2026) Physiopathology of fibrinolysis in sepsis-induced disseminated intravascular coagulation: Emerging mechanisms and pharmacological targets. Ann Intensive Care 16: 100008

13. Barrett CD, Moore HB, Moore EE, Wang J, Hajizadeh N, Biffl WL, Lottenberg L, Patel PR, Truitt MS, McIntyre RC, Jr., Bull TM, Ammons LA, Ghasabyan A, Chandler J, Douglas IS, Schmidt EP, Moore PK, Wright FL, Ramdeo R, Borrego R, Rueda M, Dhupa A, McCaul DS, Dandan T, Sarkar PK, Khan B, Sreevidya C, McDaniel C, Grossman Verner HM, Pearcy C, Anez-Bustillos L, Baedorf-Kassis EN, Jhunjhunwala R, Shaefi S, Capers K, Banner-Goodspeed V, Talmor DS, Sauaia A, Yaffe MB, (2022) Study of Alteplase for Respiratory Failure in SARS-CoV-2 COVID-19: A Vanguard Multicenter, Rapidly Adaptive, Pragmatic, Randomized Controlled Trial. Chest 161: 710–727

14. Hingorani AD, Windt DA, Riley RD, Abrams K, Moons KG, Steyerberg EW, Schroter S, Sauerbrei W, Altman DG, Hemingway H, Group P, (2013) Prognosis research strategy (PROGRESS) 4: stratified medicine research. BMJ 346: e5793

15. von Elm E, Altman DG, Egger M, Pocock SJ, Gotzsche PC, Vandenbroucke JP, Initiative S, (2007) The Strengthening the Reporting of Observational Studies in Epidemiology (STROBE) statement: guidelines for reporting observational studies. Ann Intern Med 147: 573–577

16. Singer M, Deutschman CS, Seymour CW, Shankar-Hari M, Annane D, Bauer M, Bellomo R, Bernard GR, Chiche JD, Coopersmith CM, Hotchkiss RS, Levy MM, Marshall JC, Martin GS, Opal SM, Rubenfeld GD, van der Poll T, Vincent JL, Angus DC, (2016) The Third International Consensus Definitions for Sepsis and Septic Shock (Sepsis-3). JAMA 315: 801–810

17. Iba T, Nisio MD, Levy JH, Kitamura N, Thachil J, (2017) New criteria for sepsis-induced coagulopathy (SIC) following the revised sepsis definition: a retrospective analysis of a nationwide survey. BMJ Open 7: e017046

18. Iba T, Levy JH, Maier CL, Helms J, Umemura Y, Moore H, Othman M, Thachil J, Connors JM, Levi M, Scarlatescu E, (2025) Updated definition and scoring of disseminated intravascular coagulation in 2025: communication from the ISTH SSC Subcommittee on Disseminated Intravascular Coagulation. J Thromb Haemost 23: 2356–2362

19. Shears M, Takaoka A, Rochwerg B, Bagshaw SM, Johnstone J, Holding A, Tharmalingam S, Millen T, Clarke F, Rockwood K, Li G, Thabane L, Muscedere J, Stelfox HT, Cook DJ, Canadian Critical Care Trials G, (2018) Assessing frailty in the intensive care unit: A reliability and validity study. J Crit Care 45: 197–203

20. Medcalf RL, Van den Berg E, Schleuning WD, (1988) Glucocorticoid-modulated gene expression of tissue- and urinary-type plasminogen activator and plasminogen activator inhibitor 1 and 2. J Cell Biol 106: 971–978

21. van Zaane B, Nur E, Squizzato A, Gerdes VE, Buller HR, Dekkers OM, Brandjes DP, (2010) Systematic review on the effect of glucocorticoid use on procoagulant, anti-coagulant and fibrinolytic factors. J Thromb Haemost 8: 2483–2493

22. Harrell F (2015) Regression Modeling With Applications to Linear Models, Logistic and Ordinal Regression, and Survival Analysis. Springer International Publishing AG, Switzerland

23. VanderWeele TJ, Ding P, (2017) Sensitivity Analysis in Observational Research: Introducing the E-Value. Ann Intern Med 167: 268–274

24. VanderWeele TJ, (2020) Optimal approximate conversions of odds ratios and hazard ratios to risk ratios. Biometrics 76: 746–752

25. VanderWeele TJ, (2018) On Well-defined Hypothetical Interventions in the Potential Outcomes Framework. Epidemiology 29: e24–e25

26. Pearl J, (2009) Causal inference in statistics: An overview. Statistics Surveys 3: 96–146, 151

27. Maldonado G, (2016) The role of counterfactual theory in causal reasoning. Ann Epidemiol 26: 681–682

28. Ding R, Wang Z, Lin Y, Liu B, Zhang Z, Ma X, (2018) Comparison of a new criteria for sepsis-induced coagulopathy and International Society on Thrombosis and Haemostasis disseminated intravascular coagulation score in critically ill patients with sepsis 3.0: a retrospective study. Blood Coagul Fibrinolysis 29: 551–558

29. Delabranche X, Helms J, Meziani F, (2017) Immunohaemostasis: a new view on haemostasis during sepsis. Ann Intensive Care 7: 117

30. Mavrommatis AC, Theodoridis T, Economou M, Kotanidou A, El Ali M, Christopoulou-Kokkinou V, Zakynthinos SG, (2001) Activation of the fibrinolytic system and utilization of the coagulation inhibitors in sepsis: comparison with severe sepsis and septic shock. Intensive Care Med 27: 1853–1859

31. Park KJ, Kim HJ, Hwang SC, Lee SM, Lee YH, Hahn MH, Kim SK, Lee WY, (1999) The imbalance between coagulation and fibrinolysis is related to the severity of the illness and the prognosis in sepsis. Korean J Intern Med 14: 72–77

32. Zeerleder S, Schroeder V, Hack CE, Kohler HP, Wuillemin WA, (2006) TAFI and PAI-1 levels in human sepsis. Thromb Res 118: 205–212

33. Draxler DF, Sashindranath M, Medcalf RL, (2017) Plasmin: A Modulator of Immune Function. Semin Thromb Hemost 43: 143–153

34. Keragala CB, Medcalf RL, (2021) Plasminogen: an enigmatic zymogen. Blood 137: 2881–2889

35. Cruz DBD, Helms J, Aquino LR, Stiel L, Cougourdan L, Broussard C, Chafey P, Ries-Kautt M, Meziani F, Toti F, Gaussem P, Angles-Cano E, (2019) DNA-bound elastase of neutrophil extracellular traps degrades plasminogen, reduces plasmin formation, and decreases fibrinolysis: proof of concept in septic shock plasma. FASEB J 33: 14270–14280

36. Blum MR, Tan YJ, Ioannidis JPA, (2020) Use of E-values for addressing confounding in observational studies-an empirical assessment of the literature. Int J Epidemiol 49: 1482–1494

37. Chung WT, Chung KC, (2023) The use of the E-value for sensitivity analysis. J Clin Epidemiol 163: 92–94

38. Hardaway RM, Harke H, Williams CH, (1994) Fibrinolytic agents: a new approach to the treatment of adult respiratory distress syndrome. Adv Ther 11: 43–51

39. Landoni G, Chowdary P, Meziani F, Creteur J, De Schryver N, Motsch J, Henrichmoeller I, Pages A, Peter N, Danays T, Weigand MA, Investigators T, (2024) Alteplase in COVID-19 severe hypoxemic respiratory failure: the TRISTARDS multicenter randomized trial. Ann Intensive Care 14: 170

40. Lindo CJ, Liao P, Khan A, Bhavsar D, Srivastava P, Glunz J, Fredenburgh JC, Wilmen A, Heitmeier S, Weitz JI, (2026) Antibody BAY3018250 against antiplasmin enhances plasminogen activation in vitro and thrombolysis in rabbits. J Thromb Haemost 24: 2179–2188

41. Engelmann B, Massberg S, (2013) Thrombosis as an intravascular effector of innate immunity. Nat Rev Immunol 13: 34–45

42. Gibson BHY, Wollenman CC, Moore-Lotridge SN, Keller PR, Summitt JB, Revenko AR, Flick MJ, Blackwell TS, Schoenecker JG, (2021) Plasmin drives burn-induced systemic inflammatory response syndrome. JCI Insight 6

